# Data-driven control of airborne infection risk and energy use in buildings

**DOI:** 10.1101/2023.03.19.23287460

**Authors:** Michael J. Risbeck, Alexander E. Cohen, Jonathan D. Douglas, Zhanhong Jiang, Carlo Fanone, Karen Bowes, Jim Doughty, Martin Turnbull, Louis DiBerardinis, Young M. Lee, Martin Z. Bazant

**Affiliations:** Department of Chemical Engineering, Massachusetts Institute of Technology, Cambridge, MA 02139; Department of Mathematics, Massachusetts Institute of Technology, Cambridge, MA 02139; Artificial Intelligence Group, Johnson Controls, Milwaukee, WI 53202; Department of Facilities, Engineering, and Energy Management, Massachusetts Institute of Technology, Cambridge, MA 02139; Environment, Health and Safety Office, Massachusetts Institute of Technology, Cambridge, MA 02139

## Abstract

The global devastation of the COVID-19 pandemic has led to calls for a revolution in heating, ventilation, and air conditioning (HVAC) systems to improve indoor air quality (IAQ), due to the dominant role of airborne transmission in disease spread. While simple guidelines have recently been suggested to improve IAQ mainly by increasing ventilation and filtration, this goal must be achieved in an energy-efficient and economical manner and include all air cleaning mechanisms. Here, we develop a simple protocol to directly, quantitatively, and optimally control transmission risk while minimizing energy cost. We collect a large dataset of HVAC and IAQ measurements in buildings and show how models of infectious aerosol dynamics and HVAC operation can be combined with sensor data to predict transmission risk and energy consumption. Using this data, we also verify that a simple safety guideline is able to limit transmission risk in full data-driven simulations and thus may be used to guide public health policy. Our results provide a comprehensive framework for quantitative control of transmission risk using all available air cleaning mechanisms in an indoor space while minimizing energy costs to aid in the design and automated operation of healthy, energy-efficient buildings.

## INTRODUCTION

The COVID-19 pandemic disrupted the global economy and caused the worldwide shut-down of many public and private buildings essential for daily life, including schools, gyms, religious centers, and offices [1]. At first, public health guidance focused on limiting transmission from fomites and exhaled large droplets, via surface disinfection and social distancing (such as the 6 foot rule), respectively [2]. As the pandemic continued, however, it was recognized that a dominant mode of transmission of COVID-19 is through virus-laden exhaled aerosol droplets, which are small enough to remain suspended in the air for minutes to hours and become well-mixed across indoor rooms [3, 4, 5, 6, 7, 8], so the recommended mitigation strategies shifted from social distancing to masking and improved ventilation and filtration of indoor air [9, 10, 11]. Notably, a collection of leading experts in respiratory disease transmission and building science called for a “paradigm shift” in the design and operation of indoor air control systems, analogous to historical efforts to reduce pathogen transmission through food and water sources [12], as healthy indoor air is becoming recognized as a fundamental human need [13].

In this work, we propose a physics-based, data-driven strategy to achieve this paradigm shift in healthy buildings, which integrates mathematical models of airborne disease transmission with available building data streams, and apply it to data collected from multiple buildings and indoor space types. By combining airflow measurements from building management systems (BMS) with CO_2_ concentration data and other measures of indoor air quality (IAQ) obtained from portable sensors, the underlying physics-based models are calibrated and used to simulate the transmission risk and energy consumption for each indoor space, as operated. We validate the use of simple, pseudo-steady models for airborne transmission rates and then integrate these models within a control framework to directly control for transmission rate in an energy-efficient manner. Using our workflow, public health officials can quantitatively assess mitigation strategies for indoor airborne disease transmission and recommend novel building control protocols to limit transmission while minimizing energy costs. Such quantitative analysis and automated building controls is necessary for developing healthy buildings to minimize the transmission of airborne diseases.

## METHODS

### Experiment

We collected data from diverse indoor spaces on the Massachusetts Institute of Technology (MIT) campus for several weeks in April 2022. College campuses have highly varying occupancy and a variety of room types and sizes, from large lecture halls to small offices, thereby creating an ideal environment for testing IAQ control measures. The monitored spaces are all served by heating, ventilation, and air-conditioning (HVAC) systems, which include sensors to measure and record total supply air flow, outdoor air flow (ventilation), and supply air temperature. Temporary in-room Kaiterra sensors were also deployed to collect additional measurements relevant to IAQ, including temperature, relative humidity, CO_2_ concentration, total volatile organic compounds (TVOC), PM10, and PM2.5. Only the first three measurements are used in this study, but the entire dataset is publicly available (SI 6). To measure outdoor temperature and humidity, which are relevant for estimating the energy consumption associated with ventilation, a QuantAQ sensor was placed on the roof of one of the monitored buildings. This sensor also provides size-resolved measurements of particulate concentrations, but those data streams were not used in this study. In addition, we collected information about each room, including floor area, ceiling height, use case, design occupancy, and HVAC configuration (SI 1).

The overall workflow for this study is illustrated in Fig. 1. The key idea is that all data streams are integrated with physics-based models to predict and control transmission risk and energy consumption. This data fusion ensures that all space-specific variables in the model are known or accurately estimated, thus allowing different spaces to be compared with confidence. Example time series data for four of the monitored rooms are shown in Fig. 2 and exhibit variations on the scales of days, hours, and minutes. The CO_2_ and humidity measurements come from the in-zone IAQ sensors, while outdoor-air flow measurements come from the BMS or are estimated from CO_2_ measurements. Of these measurements, CO_2_ concentration is the most strongly varying, as it is driven primarily by room occupancy. The peak values for Classroom 3 are significantly higher than for the other rooms, as it is does not have a forced supply of outdoor air provided by the HVAC system and is thus only naturally ventilated.

**Figure 1:**
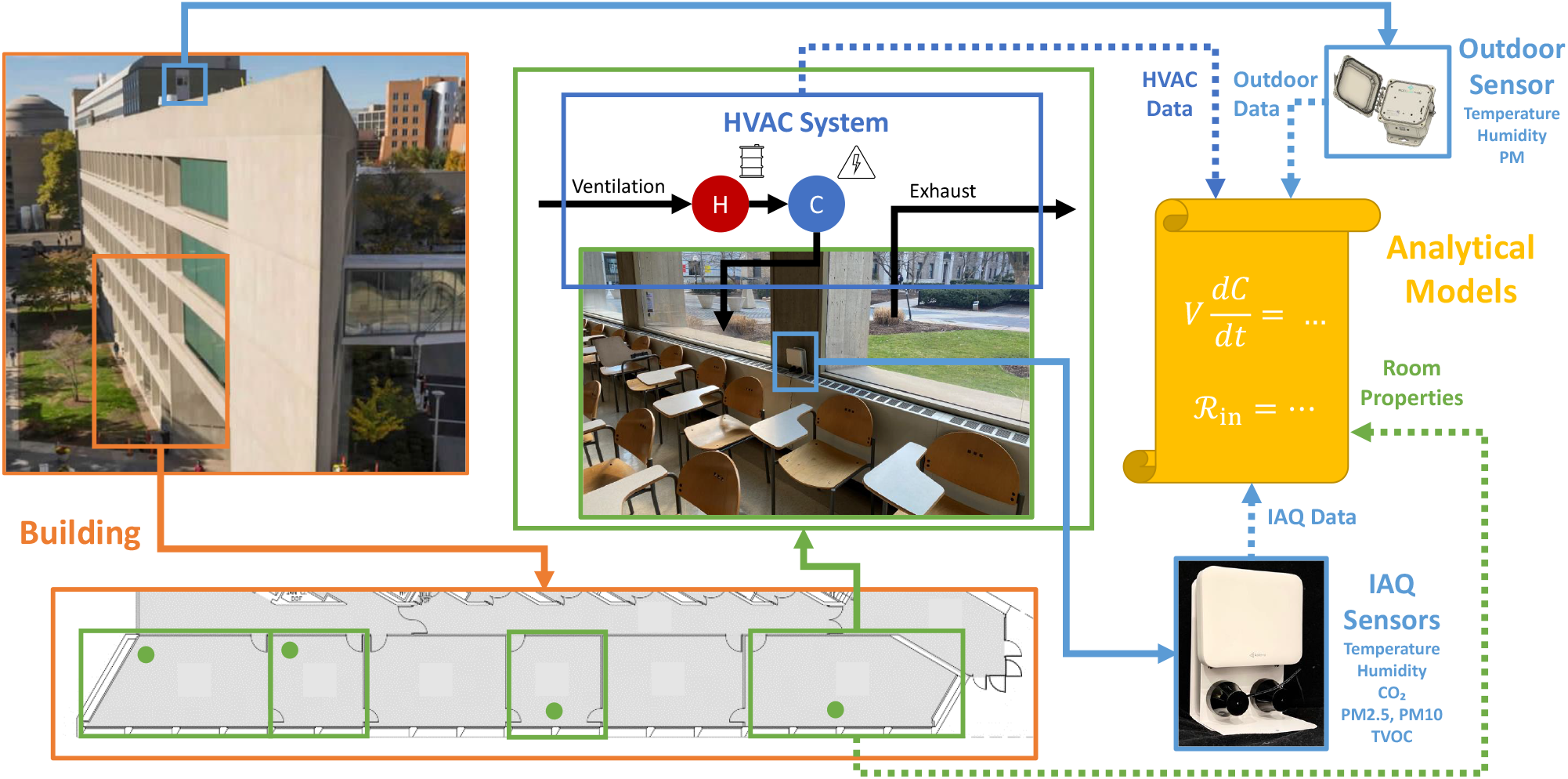
Diagram of overall workflow. Portable indoor air quality (IAQ) sensors are placed in each monitored room, and a more durable sensor is placed on the roof to record outdoor conditions. These sensor measurements are then combined with time-series data from the heating, ventilation, and air conditioning (HVAC) system and basic properties of each room (area, ceiling height, design occupancy, etc.) for analysis.

**Figure 2:**
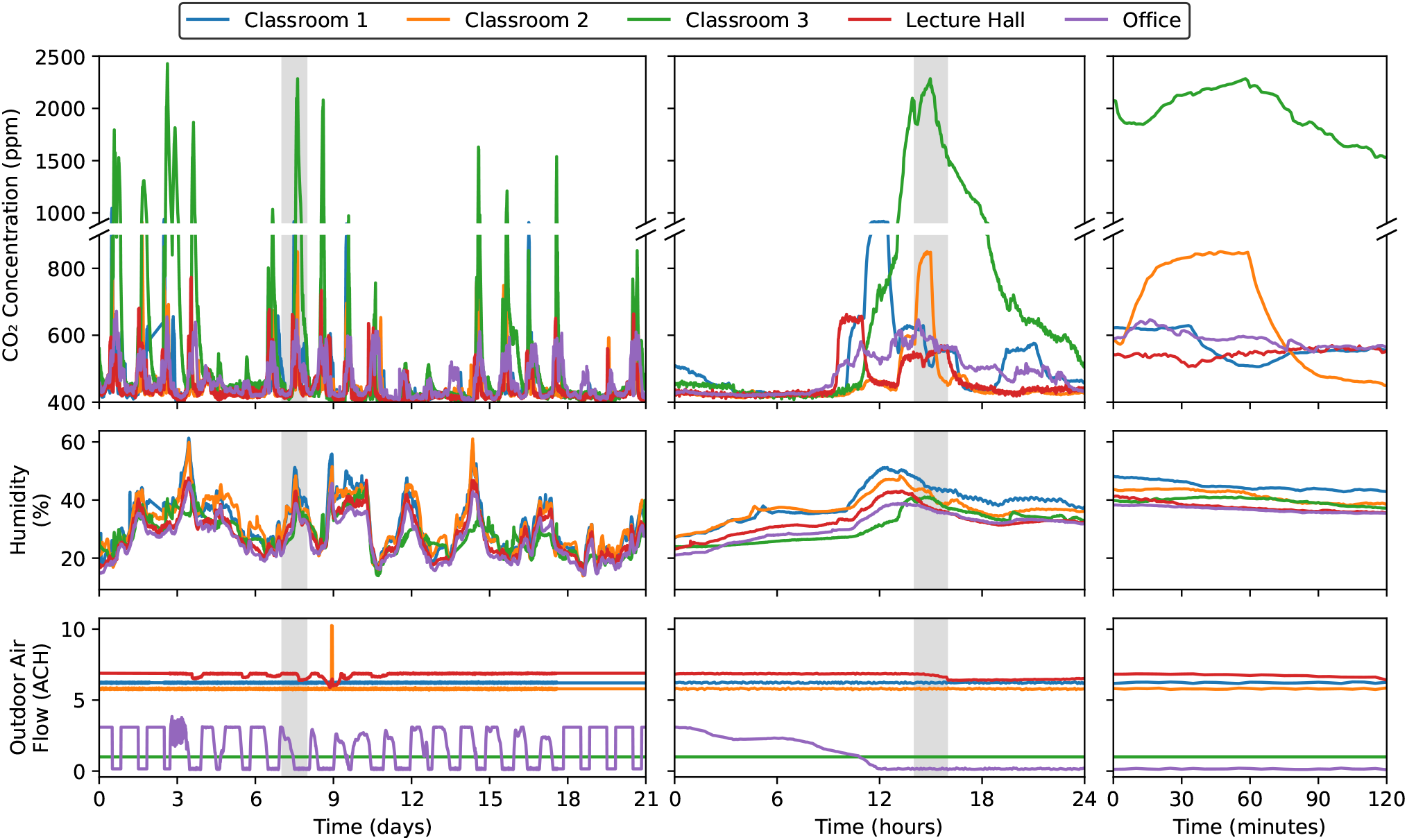
Plot of sample time series data from the study period at multiple time scales (days, hours, and minutes). The shaded region in the first (or second) column shows the time range covered in the second (or third) column. Missing data points have been filled in via interpolation.

### Theory

#### Infectious particle model

Simple mass-balance models [14, 15] have been used for decades to study airborne transmission via aerosols and have been successfully applied to explain transmission in prior diseases [16, 17] and COVID-19 [18, 19, 20, 9, 21]. Each room is assumed to be well-mixed, such that the particle concentration can be treated as uniform throughout the room [14, 22]. This assumption is shown to produce high-quality occupancy estimates, indicating sufficient accuracy for our purposes. The mass balance for infectious particles in a room results in the following partial differential equation model for the time-evolution of infectious pathogen concentration, *C* (*r, t*), per droplet size in a room of volume *V* and area *A* [9]:

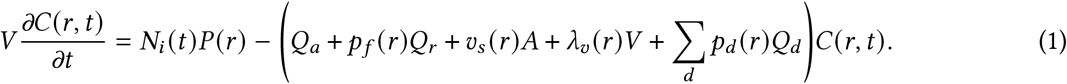

where *N*_*i*_ are the number of infectors present in the room exhaling infectious droplets with rate *P* (SI 2). Infectious droplets are removed through ventilation (*Q*_*a*_), filtration in the recirculated airflow (*pf* (*r*)*Q*_*r*_), sedimentation (*v*_s_ (*r*)*A*), deactivation (*λ*_v_ (*r*)*V*), and the action of disinfection devices (∑_*d*_ *p* (*r*)*Q*). All removal mechanisms can be expressed as rates, *λ*_*a*_ = *Q*_*a*_/*V, λ*_*f*_ (*r*) = *pf* (*r*)*Q*_*r*_ /*V, λ*_*s*_ (*r*) = *v*_*s*_ (*r*)*A*/*V*, and *λ*_*d*_ (*r*) = ∑_*d*_ *p*_*d*_ (*r*)*Q*_*d*_ /*V*, and lumped into a single parameter the describes the supply of “equivalent outdoor air” (EOA) delivered to the space, *λ*_EOA_ = *λ*_*a*_ +*λ*_*f*_ +*λ*_*s*_ +*λ*_*v*_ +*λ*_*d*_ [23]. EOA quantifies each removal mechanism in terms of volumetric flow of outdoor-air ventilation that would lead to an equivalent removal rate of infectious particles, thus facilitating comparisons among disparate processes.

#### Safety Guideline

Following [9], the model can also be approximated analytically to derive a “safety guideline” that bounds the indoor reproductive number, ℛ _*in*_, defined as the expected number of transmissions if an infector were present for a time *τ* in a given room:

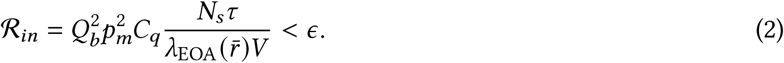

where *C*_*q*_ represents the infectious quanta concentration in exhaled air (SI 3); *Q*_*b*_ is the occupants’ breathing rate; *p*_*m*_ is a mask penetration factor of aerosols; *V* is the volume of the room; *N*_*s*_ is the number of susceptible occupants; and *ϵ* is the desired risk tolerance.

Droplet size dependencies are integrated out by defining an effective droplet size 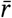 (SI 3). Typical ranges for these parameters and the values used in this study are provided in Supporting Information (SI 7).

Simply put, ℛ _*in*_ is proportional to the product of susceptible occupants and the time spent, divided by the EOA provided to the room. Thus, any holistic risk assessment and transmission mitigation strategy must consider all three dimensions. For example, simply mandating a maximum occupancy in an indoor space may not adequately reduce transmissions if the occupants spend a large amount of time in the space. Alternatively, occupancy limits may be unnecessary, if an appropriate amount of EOA is delivered to the space.

#### CO_2_-based Safety Guideline

CO_2_ is often used as an indicator of transmission risk [24, 25, 26, 27], but we stress that it is only a partial proxy that requires care to interpret, especially when comparing across spaces with significantly different HVAC systems. In particular, the dynamical model for CO_2_ concentration, 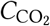, is

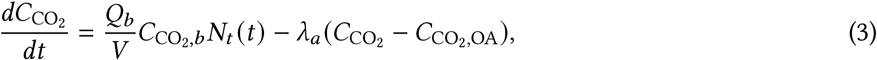

where *Q*_*b*_ is the occupant breathing rate, 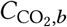 is the exhaled-breath excess CO_2_ concentration, and 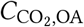 is the outdoor-air CO_2_ concentration. To derive a CO_2_-based guideline, a pseudo-steady analysis of the dynamical model for CO_2_ concentration can be combined with Equation (2) [25], which incorporates the differences between infectious particle dynamics and CO_2_ dynamics stemming from sources of EOA beyond ventilation. We thus arrive at the following safety guideline based on CO_2_ concentration measurements:

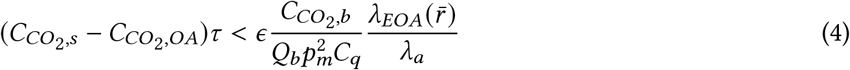

#### Short-Range Transmission

Short-range respiratory flows also contribute to the risk of disease transmission in indoor spaces [28, 29, 30] and should be compared with the risk of long-range airborne transmission in any safety guideline [9].

Estimates of short-range transmission rates can be derived from the theory of turbulent jets [9]. This analysis predicts that the concentration of infectious particles in the jets of infectors’ exhaled breath decays as 1/*x* where *x* is horizontal distance. A key deficiency of this model is that it does not account for the buoyancy of exhaled breath, which causes it to quickly rise out of the breathing zone of a potential susceptible. Thus, rather than use the turbulent jet models directly, we instead employ an empirical model derived from the experimental results of [31]. In this paper, Zhang *et. al*. calculate a “susceptibility index” defined as *ϵ* = *C* (*x*)/*C*_∞_ where *C* (*x*) is the infectious particle concentration at horizontal distance *x* from the mouth of the infector, and*C*_∞_ is the background room concentration. For our purposes, we assume the concentration within the jet follows the model

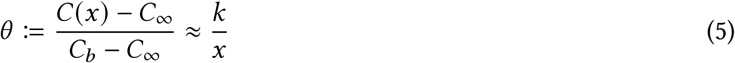

where *k* is an unknown constant to be determined. Assuming *C*_∞_ = *C*_*b*_*Q*_*b*_/*V λ*_EOA_ follows the pseudo-steady well-mixed model, we can derive the relationship

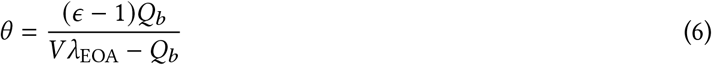

To quantify the short-range transmission risk, we use the model

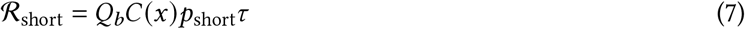

in which the new parameter *p*_short_ represents the probability that a susceptible is directly within the short-range plume exhaled by each infector. Note the similar form to the well-mixed formulation (2), with the primary difference that the short-range risk depends on occupant separation distance *x*. More information about how we determine this parameter is provided in SI 4.

#### Occupancy and Ventilation Estimation

A key factor affecting the transmission rate is the time-varying occupancy in each space, which is then used to estimate the numbers of susceptible, *N*_*s*_, and infectious, *N*_*i*_, occupants. There are various occupant-counting technologies available, which utilize images, sound, or IAQ data, such as CO_2_ concentration, temperature, and humidity [32, 33, 34, 35, 36, 37, 38, 39, 40, 41, 42, 43, 44, 45]. Depending on the data that the method uses, the required equipment may be expensive and complicated to install, so many spaces will not have occupancy counts directly available. In addition, many of these technologies may compromise the privacy of occupants by using video or images of occupants. Finally, machine learning methods which utilize multiple streams of environmental data often require individual training data for each room, which is infeasible to apply to a large number of rooms. One alternative would be to simply assume a fixed occupancy count and time-varying schedule (e.g., as provided by [46]) for each space, but this approach would likely introduce unacceptably high error, especially when occupancy is highly variable. Given these challenges, we thus estimate occupancy by modifying previous methods that solve the full inverse problem of the CO_2_ concentration dynamical model given by Eq. (3) and data, with a novel extension to simultaneously estimate ventilation rates if not measured. This method protects occupant privacy and requires no training data, so it can be easily implemented in new spaces.

When the ventilation rate *λ*_*a*_ (*t*) is known (e.g., due to direct measurement, which is often the case for spaces with modern HVAC systems), we can estimate occupancy *N*_*t*_ (*t*) by choosing a set of basis functions and finding the linear combination of those basis functions such that the predicted time series of 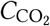 under (3) matches the actual measured values as closely as possible. This step is performed by embedding a discretized version of this model inside an optimization problem and solving for the basis-function coefficients via optimization techniques. Mathematically, we denote the basis functions *ϕ*_*i*_ [*t*] for *N*_*t*_. To embed the model, we choose a fixed sample rate Δ = 1 min and define a new function 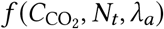 to give the explicit Runge-Kutta 4 discretization of the ODE model (3). The resulting optimization problem is thus

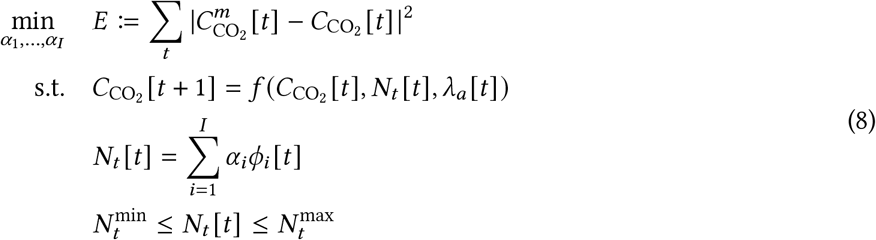

given CO_2_ concentration measurements 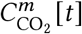 and pre-defined occupancy bounds 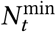 and 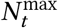 (which we set respectively to zero and 1.5 times each room’s design occupancy). Note that we use square brackets to emphasize that these quantities are defined in *discrete* time. Because the function *f* (·, ·, ·) is *linear* in its first and second variables, the resulting optimization problem is thus a convex quadratic programming problem that can be solved using standard techniques.

In cases where the ventilation rate *λ*_*a*_ (*t*) is *not* measured or otherwise known, the proposed strategy requires some modification. One possible method to estimate its value would be to add corresponding basis functions for *λ*_*a*_ [*t*] and embed them in the optimization problem with new decision variables analogous to the treatment of *N*_*t*_. For spaces with sufficiently low ventilation rates (and sufficiently high sample rate for CO_2_ concentration measurements), this modification can be sufficient, with the resulting ventilation and occupancy estimates being the ones that most closely match the measured CO_2_ data. However, this change would render the resulting optimization problem nonconvex, and selection of appropriate basis functions could be challenging. In addition, if there are few changes in occupancy throughout the day, the chosen objective function is dominated by pseudosteady periods with 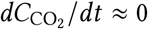, which thus implies the degenerate relationship *λ*_*a*_ ∝ *N*_*t*_ and creates additional problems as discussed below. Therefore, we instead opt for a slightly different approach.

Assuming the ventilation rate *λ*_*a*_ (*t*) is *constant* 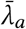, over the time horizon, we let 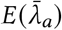 denote the optimal value of the optimization problem (8) assuming 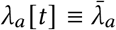, and we let 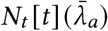 denote the corresponding value of the occupancy profile. To estimate 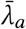, we can thus solve the one-dimensional optimization problem

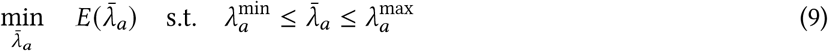

by using the volume-normalized *λ*_*a*_ as the decision variable, as we know its value should almost always be between 1 and 10 h^-1^ independent of the specific room. Thus, despite its nonconvexity the problem can be solved by simple bounded scalar optimization techniques, or even via an exhaustive grid search with a chosen granularity. To estimate uncertainty in the estimate we take a level set of the objective function, with the threshold set to 50% higher than the optimal value 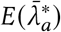.

If there are a sufficient number of large occupancy changes, this procedure can produce a tight range for the estimated ventilation, primarily by matching the exponential decay predicted by the model during such events to the measured data. However, if occupancy is relatively constant, the data will be dominated by the pseudo-steady relationship

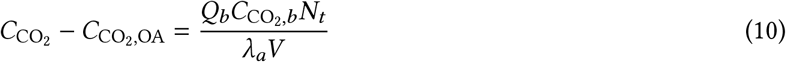

which is linearly degenerate, so the resulting uncertainty region will be extremely large.

To break this degeneracy, we note that while we certainly do not know the full time-varying occupancy profile, we often have a good idea of *peak* occupancy 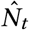 over the given time period. Letting 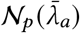 denote the *p*th percentile of the occupancy estimates *N*_*t*_ [*t*] (*λ*_*a*_), we thus desire that 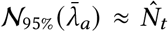. (We use the 95th percentile to add some degree of robustness to small periods of abnormal data, e.g., when the HVAC system is shut down for maintenance.) Adding this relationship to the cost function, we thus arrive at our final modified optimization problem

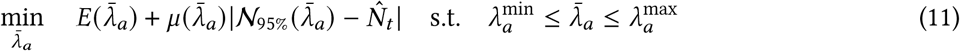

in which *μ* (·) is a scaling factor to weight the two terms. We use

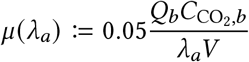

so that this penalty accounts for (5% of) the error that would be induced in the pseudo-steady model due to the difference in occupancy. In practical applications, this scale factor would be adjusted up or down depending on confidence in the assumed peak occupancy 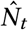. For each room, the assumed value of 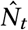 is generally set equal to 50-100% of design occupancy consistent with typical usage during the monitoring period.

#### Transmission-Controlled Ventilation

While addressing public health concerns, there are still many opportunities to reduce energy consumption in buildings, which account for 18% of total energy use in the United States [47]. Long term building operation must balance airborne transmission risk and IAQ with energy consumption.

Formulating all removal processes in terms of EOA provides a common basis to compare various technologies in terms of cost per ACH of EOA. Under this lens, filtration, which can either be provided by in-room air cleaners or recirculated supply air, is often a much more energy-efficient source of EOA than ventilation. As an alternative, ultraviolet (UV) light can provide significant EOA by eradicating any infectious material within particles [48, 49, 50, 51] and can be installed in an upper-room configuration with shielding or as “far-UV” that is not harmful to occupants [52]. If properly installed, such systems can deliver EOA even more efficiently than filtration-based sources [53]. Thus, to optimize energy efficiency, it is necessary to consider all of these options.

Unfortunately, the primary source of EOA for many rooms is in fact ventilation. Considering the large variance in transmission risk for spaces with strongly time-varying occupancy, however, it is possible to significantly reduce energy costs by limiting extra ventilation to periods of high occupancy. Here, we analyze and compare demand-controlled ventilation (DCV) and transmission-controlled ventilation (TCV) operation modes. DCV is a feedback control mechanism implemented in most modern HVAC systems, which adjusts ventilation rates in real time to maintain a setpoint of CO_2_ concentration. This method thus does not consider any other EOA sources, and thus the mapping from CO_2_ setpoint to transmission risk can vary strongly from space to space. However, given that our primary goal is to control the transmission risk in each room, we propose TCV as a novel operating mode to maintain a transmission-rate setpoint by interfacing with HVAC and accounting for other sources of EOA, all of which impact airborne disease transmission.

To implement this control strategy, we first need to evaluate the current transmission rate 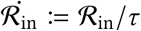. The pseudo-steady model gives 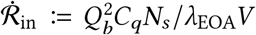. The value of *λ*_EOA_*V* can be calculated using flow measurements and filtration parameters for the BMS-provided clean air and the humidity measurements and physics-based models for the deposition and deactivation components of EOA. To avoid the need for an explicit estimate of *N*_*s*_, we define the CO_2_ generation rate as 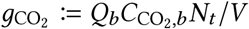 and make the (conservative) assumption that *N*_*s*_ ≈ *N*_*t*_. We thus arrive at the formula

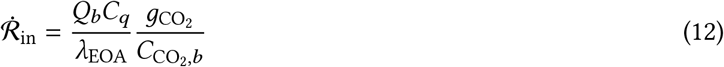

which can be evaluated by the BMS. The primary benefit is that 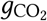 can be estimated directly from successive measurements of 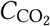 and *λ*_*a*_ in accordance with the dynamic model (3).

To define the action of the controller, we thus take a transmission-rate setpoint 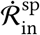 (chosen in accordance with expected exposure time *τ*) and invert the previous formula to find the corresponding EOA setpoint

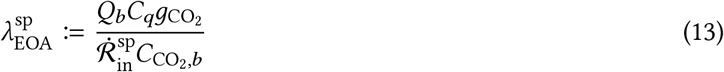

From this value, the BMS can adjust its various setpoints to deliver the required amount of EOA. In cases where the BMS can control multiple sources of EOA (e.g., ventilation, filtration via recirculation, and possibly in-zone disinfection devices), some form of prioritization would be needed, for example selecting in order of increasing energy consumption. More information on the corresponding control logic is provided in SI 5.

## RESULTS

### Occupancy and ventilation estimation

To validate the proposed approach for occupancy estimation, we manually collected a limited amount of occupancy data in two rooms. In Classroom 1, attendance was taken at each class, which was assumed to be constant throughout the 90-minute lecture period. In Office, a sign-in/sign-out sheet was used over a three day period to estimate time-varying occupancy. Fig. 3 shows the estimated time-varying occupancy in both rooms throughout the monitoring period, as determined by the solution to (8). During nominal occupied hours, we see that the estimates are in good agreement with measured occupancy where available. In addition, we note the strong time-varying character of these curves, which emphasizes the need to use realistic occupancy profiles (rather than simple fixed schedules) to accurately assess transmission risk for these spaces.

**Figure 3:**
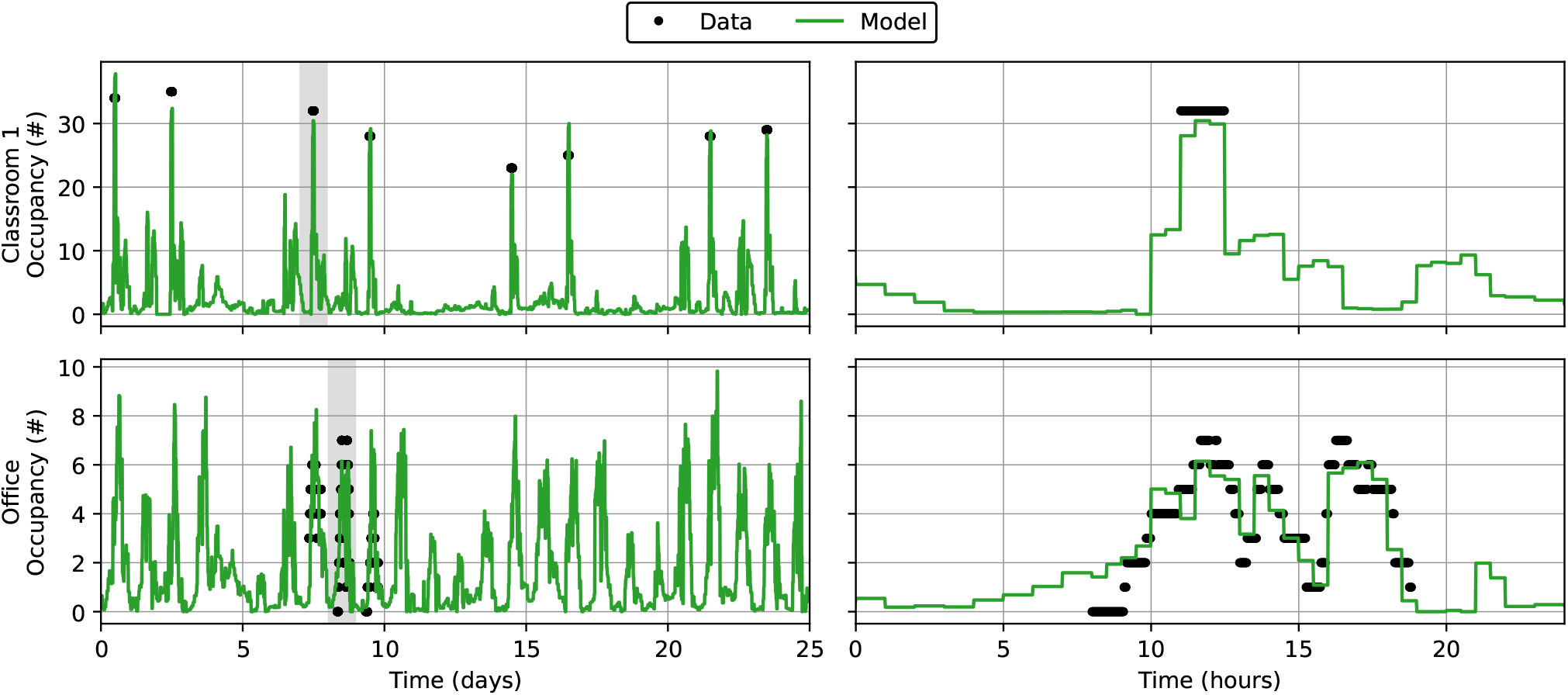
Fit occupancy profiles for rooms with (partial) occupancy data. Occupancy is assumed to be constant over each 30-minute interval in the optimization formulation. Despite simplicity of the well-mixed models, estimated occupancy profiles closely match the available occupancy data.

To illustrate the proposed approach for simultaneous occupancy and ventilation estimation, Fig. 4 shows the objective functions, model fits, and estimated occupancy profiles for a 1-day period in Classroom 1. Note that the grey “CO_2_ Fit” objective corresponds to the formulation in (9), while the black “+Occupancy Penalty” is the modified formulation in (11). As mentioned before, uncertainty regions are calculated as ±50% of the optimal objective value. Including only the penalty on CO_2_ concentration fit, we see that the estimated ventilation rate is quite low, with a large relative uncertainty. Although the CO_2_ error rules out the extremely low and high ventilation rates, it cannot adequately distinguish between the intermediate values. However, after adding the additional term for deviation from the peak occupancy target (set to 65% of the room’s design occupancy), the estimated value is now much closer to the actual measured value, with lower relative uncertainty.

**Figure 4:**
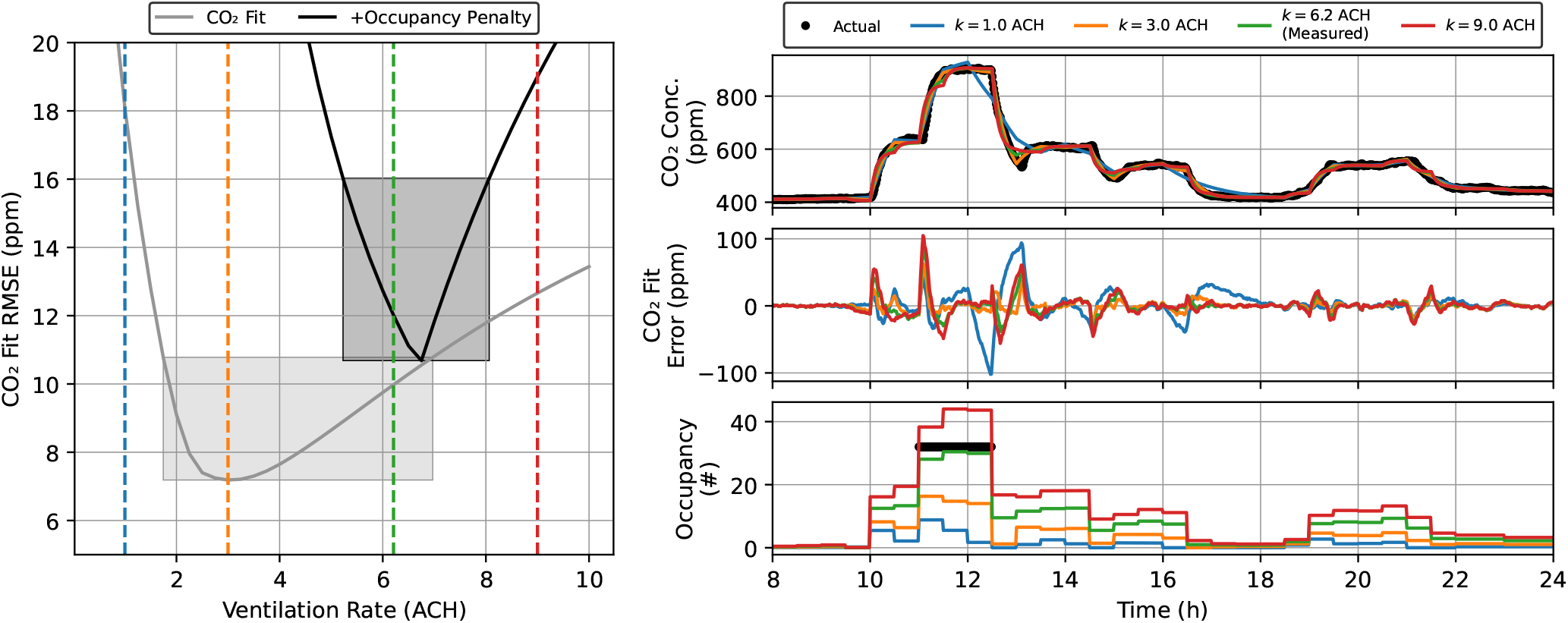
Simultaneous estimation of ventilation rate and time-varying occupancy. Left: objective function with (black) and without (gray) the peak-occupancy penalty along with uncertainty regions. The optimization procedure chooses the ventilation rate with the lowest value of these objective functions. Right: simulated CO_2_ concentrations, fit errors, and estimated occupancy for selected ventilation rates. These values correspond to the dashed colored lines on the left. Note that the measured ventilation rate for this room was 6.2 ACH, which corresponds to the green curves.

### Infection risk

The workflow can be assessed by calculating the transmission risk in the different indoor spaces using the collected data along with the full dynamical model and the pseudo-steady approximation. The transmission rate values predicted by the full model and pseudo-steady approximation are in excellent agreement, and where they differ, the pseudo-steady model produces more conservative estimates (Fig. 5). We assess these models in two ways. In Fig. 5, we extract random segments of data from Classroom 2 (during nominally occupied hours) and plot points for each segment on a plane with axes for average occupancy and time. The color of each point corresponds to the event reproductive number calculated from the full dynamical model. Since EOA delivery is essentially constant for this space, the pseudo-steady model predicts that ℛ _*in*_ ∝ Occupancy × Time, which is the same trend exhibited by the color of the points in Fig. 5. This validates the use of the safety guideline, Eq. (2), to limit occupancy and time such that ℛ _*in*_ is below a given tolerance, as indicated by the dashed curve, which can be shifted by altering the amount of EOA provided in the room.

**Figure 5:**
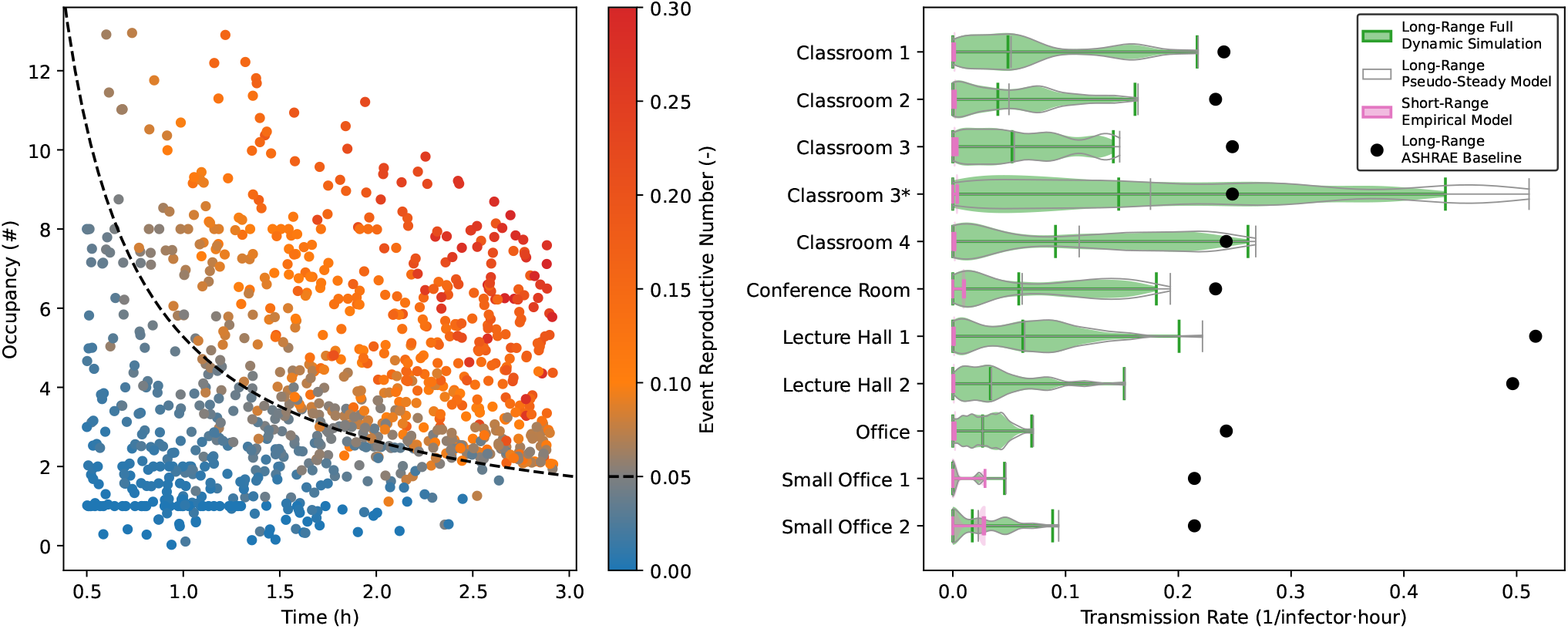
Transmission predictions for selected rooms in the study. Left: Scatter plot of the reproductive number for randomly chosen time periods from the full dynamical simulations, found to be in good agreement with the safety guideline [9] from the pseudo-steady formula, Eq. (2). Right: Distributions of transmission rates throughout the study period computed using full model simulation and pseudo-steady approximation, again showing good agreement. The distributions are weighted by occupancy and thus predict the expected number of transmissions if one occupant were to be infectious for one hour. Green curves are the distributions from the full dynamic model, with green lines showing the minimum, median, and maximum values. Gray curves are the distributions from the pseudo-steady model, with grey lines showing the same three statistics. Black dots show expected values assuming minimum ventilation rates and occupant density per ASHRAE standard 62.1 [55].

To assess other spaces, Fig. 5 shows distributions of transmission rates across all the monitored spaces. These points are based on a 5-minute sample rate for both models, and the distributions are weighted by the number of occupants within each point. The black dots indicate the corresponding steady-state transmission rate for a space of that type with baseline ventilation rates and occupant density, per ASHRAE standards. We see that, in almost all spaces, the worst-case transmission rate is below the baseline value as expected, since MIT buildings were deliberately operated with extra ventilation during the monitoring period to limit COVID-19 transmission. Again, the distribution of transmission rates calculated from the pseudo-steady model (gray curves) closely matches the distribution of transmission rates calculated from the full dynamic model (green curves). The median values of these distributions are generally within 1% agreement, and the maximum values differ by less that 10%.

The primary outlier from this trend is Classroom 3*, which has no mechanical ventilation, resulting in significantly smaller EOA delivery than in the other spaces. As a result, when a large number of people enter this room, the infectious particle concentration takes longer to approach the pseudo-steady values. As such, the pseudo-steady model predicts a more conservative, higher transmission rate. When a large number of people leave those spaces, the opposite transient effect occurs, but since the occupancy is generally lower as people are leaving the space, those events are weighted less in the final distribution. The net result is that the pseudo-steady model predictions are slightly conservative.

The results shown for Classrooms 3 and 3* are for the same space, occupancy profile, and ventilation rate, but different levels of filtration. We assume an active filter delivering 4.5 ACH of HEPA filtration in Classroom 3 and inactive filter in Classrom 3*. When the filter is inactive, there is roughly a threefold increase in median transmission rate. We highlight this distinction because measured CO_2_ concentrations would be exactly the same for the two scenarios, since filtration is a form of EOA that does not impact CO_2_, which thus cannot be used by itself to assess transmission risk. Instead, safety guidelines [9, 25], which incorporate the differences between total EOA and ventilation, based on a fixed ℛ _*in*_ tolerance, are more appropriate to account for differences in indoor spaces.

We also conservatively estimate that for most of the spaces monitored, short-range effects (pink curves) account for less than 5% of the expected transmissions caused by long-range mixing using Eq. (7). The only exception is for rooms where close-range face-to-face contact between occupants is common, such as in the Small Office spaces, where short-range risk may reach up to 30% of the long-range risk. However, the flows responsible for short-range transmission can be eliminated by requiring occupants to wear masks [54].

### Energy and control analysis

Given a desired level of total transmission risk, buildings should operate to achieve that risk as efficiently as possible, taking advantage of all available mechanisms of infectious particle mitigation. Here, we analyze and compare DCV and TCV, our novel control strategy, operation modes.

To quantitatively assess the inherent tradeoff between energy consumption and transmission risk, we estimate the daily energy cost for each room as operated and under various hypothetical scenarios using standard thermodynamic and equipment modeling procedures from our previous work [56, 57]. For the monitored rooms, the primary cost driver is the energy required to heat the outdoor air up to its supply temperature.

We refer to the actual operation during the monitoring period as the “Baseline” scenario. The hypothetical scenarios considered for each room are as follows:

- Curtailed: ventilation is supplied at the same rate as observed in the data only during nominal occupied hours, assumed to be 8 am through 10 pm.
- In-Zone Filtration: in addition to the schedule change in the “Curtailed” scenario, 2 ACH of in-room filtration is provided via standalone air cleaners active during occupied hours.
- In-Zone Far UV: in addition to the schedule change in the “Curtailed” scenario, upper-room far UVC lamps are activated during occupied hours so as to provide 5 ACH of EOA.
- ASHRAE Minimum: ventilation follows the “Curtailed” schedule and is further adjusted to provide the minimum amount of ventilation required in each space per ASHRAE standard 62.1 [55].
- Demand Controlled (DCV): ventilation is provided by a standard demand control algorithm for a given CO_2_ concentration setpoint.
- Transmission Controlled (TCV): ventilation is provided by a modified algorithm that provides enough ventilation to operate below a given transmission risk as calculated by the pseudo-steady model.

After estimating the time-varying ventilation that would be provided by each hypothetical strategy, transmission risk and energy cost can be calculated using the modeling approach discussed previously. These results are shown for three representative spaces in Fig. 6. Similar plots for other spaces are provided in the Supporting Information.

**Figure 6:**
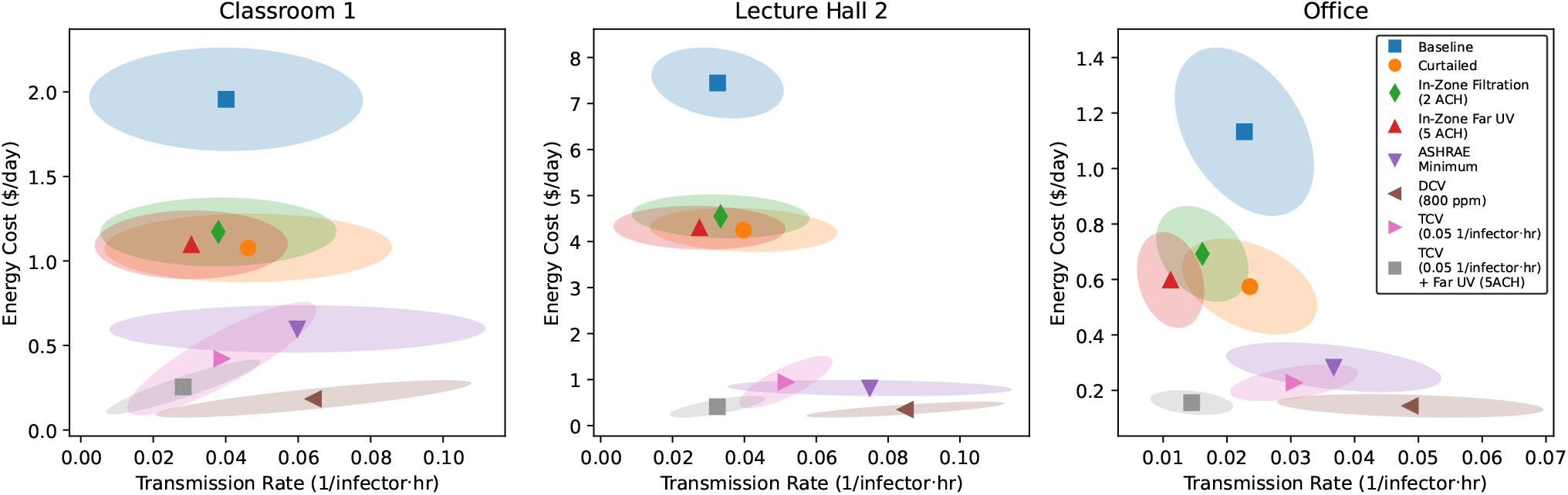
Summary of energy versus transmission rate tradeoffs for hypothetical ventilation scenarios. Points and shaded regions show means and joint standard deviations for daily values within the study period.

In all spaces in Fig. 6, we see there are significant opportunities to reduce energy consumption without large changes in the average and spread of the transmission rate. Simply curtailing ventilation during nighttime unoccupied hours cuts energy consumption roughly in half, with only a slight increase in transmission rate due to a small number of after-hours gatherings in that room. Adding in-room filtration can reduce transmission below baseline values while still providing significant reduction in energy cost. Based on current experimental data [52], far UV disinfection is even cheaper than in-room filtration. The ASHRAE minimum protocol further reduces energy consumption but increases the mean and spread of the transmission rate. However, by applying some of the more advanced control algorithms, transmission rate can be maintained near or below a desired threshold while maximizing energy savings. In particular, DCV at 800 ppm achieves minimum energy cost with transmission risk, while the novel TCV strategy at 0.05 per infector·h forgoes some of the energy savings to achieve further reduction in observed transmission rate. Note that the specific setpoints of these two strategies could be adjusted up or down to further tune the tradeoff. Finally, combining TCV with in-room far UV can achieve the same average and lower spread of transmission rate as the conservative baseline schedule with up to a tenfold decrease in energy cost.

Overall, these results illustrate that advanced control strategies and alternative sources of EOA can be employed to provide similar or better expected transmission rate while significantly reducing energy costs compared to constantly operating at high ventilation rates. We see that the TCV strategies deliver Pareto-optimal performance, which demonstrates that the pseudo-steady transmission model is sufficiently accurate to achieve its control objectives while remaining mathematically simple enough to integrate into existing HVAC control logic. Such TCV systems could interface with other sources of EOA, such as filtration and UV disinfection, to simultaneously control transmission rates and minimize energy consumption by prioritizing lower energy sources.

## CONCLUSIONS

Our framework successfully combines data-streams from sensors with accurate physical models of aerosol physics and disease transmission to predict transmission rates in real-life, as-operated indoor spaces. Such an approach can be used both for real-time transmission control and future building design when considering indoor air quality. We have demonstrated the possibility of transmission-controlled ventilation by implementing our models in HVAC control logic, which maintains air quality at a safe level while optimally minimizing energy usage. Our simple formulas provide an easy-to-use design framework for building designers and engineers when considering the trade-off between energy efficiency and indoor air quality for installing various clean air delivary mechanisms in current and future buildings.

Our results can also inform public health guidance, using data from real buildings. We have validated the use of the simple safety guideline, Eq. (2), to limit infection risk in different classes of indoor spaces, rather than strict occupancy limits [9]. We have shown that the underlying pseudo-steady approximation is consistent with full, dynamical simulations, and whenever small discrepancies arise, the guideline always provides a more conservative estimate of the risk. For normal occupancy in the monitored spaces, we also predict that short-range transmission via respiratory flows can be neglected (compared to the long-range airborne transmission) without imposing physical distance limits.

CO_2_ measurements play a central role in the analysis. Time-varying occupancy and ventilation rates are critical parameters in the models for transmission rate. When rooms are mechanically ventilated with set rates from the BMS, CO_2_ measurements can be used to accurately estimate occupancy with complete anonymity to the occupants. When ventilation rates are unknown, such as for naturally ventilated spaces, CO_2_ measurements can be used to estimate both occupancy and ventilation rates, albeit with larger uncertainty. Importantly, we demonstrated how CO_2_ is not a direct proxy for transmission risk due to varying sources of EOA. However, with appropriate knowledge of the EOA sources, CO_2_ can be used in conjunction with our models to accurately estimate transmission risk in diverse indoor spaces.

Controlling disease transmission must also be considered within the context of broader societal needs, such as minimizing energy usage, pollution, and carbon emissions. Our framework is able to quantify and optimize these tradeoffs, as the various sources of EOA are all incorporated, including their different energy requirements. We demonstrate how this modeling can be used to design optimal control protocols which control for certain transmission risk setpoints while also minimizing energy requirements by prioritizing low-energy EOA sources. Enabling real-time control of infection risk, prioritized against energy consumption, is a critical first step in the paradigm shift toward more healthy, energy efficient buildings [12].

## Data Availability

All data produced are available online at https://github.com/acoh64/MIT-JCI-IAQ-HVAC

## ACKNOWLEDGEMENTS

A.E.C. was supported by the Department of Defense (DoD) through the National Defense Science and Engineering Graduate (NDSEG) Fellowship Program. Partial funding for this study was provided by Johnson Controls, and portions of the research are subject to patents pending.

## AUTHOR CONTRIBUTIONS

^*^ M. J. R. and A. E. C. contributed equally to this work.

M.Z.B. and Y.M.L. conceived the study. M.J.R. wrote the code and performed the simulations. M.J.R., J.D.D., A.E.C., Z.J., C.F., K.B., J.D., M.T., and L.B. set up the sensors and planned experiments. M.J.R. and A.E.C. prepared the manuscript. All authors discussed the results and edited the manuscript.

## SUPPLEMENTARY INFORMATION

Supplementary Information is available for this work.

## DATA AVAILABILITY STATEMENT

All data used in this study is available at https://github.com/acoh64/MIT-JCI-IAQ-HVAC.

