## Supplementary Information for "Data-driven control of airborne infection risk and energy use in buildings"

April 9, 2023

#### Contents

**1 Room Details** **2**
**2 Transmission Modeling Details** **9**
**3 Safety Guideline Derivation** **11**
**4 Short-Range Transmission Risk** **21**
**5 Hypothetical Ventilation Strategy Details** **23**

*M.J.R. and A.E.C. contributed equally to this work.

**6 Data Availability** **28**

**7 Transmission Parameter Values** **31**

### 1 Room Details

The rooms monitored in this study contain a dedicated-outdoor air HVAC configuration. In a dedicated-outdoor air setup, a dedicated stream of pure outdoor air is delivered to each room for ventilation purposes, with a corresponding amount of room air exhausted to maintain pressure. Room temperature is maintained by in-room fan-coil units (FCUs) that use a fan to recirculate room air across heating or cooling coils. This recirculated air stream is not filtered.

In contrast to this dedicated outdoor-air setup is a "mixed-air, single-duct" configuration, which is also common in many schools and commercial buildings. With this design, the outdoor air required for ventilation is centrally mixed with (some fraction of) the return air from served spaces, after which the mixture is heated or cooled to a desired supply temperature before being split and delivered to each space in accordance with its thermal needs. For the purposes of airborne transmission, the primary difference is that the mixed-air stream is passed through a filter, and thus the recirculating fraction of the air is partially cleaned, serving as a supplemental source of EOA. Indeed, if this filter is rated MERV-13 or higher, upwards of 95% of the potentially infectious aerosols are removed from the recirculated air. Thus, for a space receiving 2 ACH of outdoor air but 5 ACH of total supply air in a mixed-air single-duct configuration, the resulting EOA delivery is almost 5 ACH. By contrast, a room in a dedicated-outdoor-air configuration would receive only the 2 ACH of ventilation as EOA.

Parameters for each monitored room are summarized in Table 1. The subsections that follow present timeseries data and energy versus transmission rate tradeoffs for each room.

Table 1: Parameters for each monitored room. ASHRAE Minimum Ventilation is calculated in CFM per standard 62.1-2019 [1] and converted to ACH via the space volume. Assumed Maximum Ventilation is calculated based on assumed design flows for each space and is used only when simulating the hypothetical ventilation scenarios.

| Name | Area ($ft^2$) | Ceiling Height (ft) | Design Occupancy (#) | Assumed Peak Occupancy (#) | Mean Ventilation (ACH) | ASHRAE Minimum Ventilation (ACH) | Assumed Maximum Ventilation (ACH) | In-Zone Filtration (ACH) |
|---|---|---|---|---|---|---|---|---|
| Classroom 1 | 734 | 10.7 | 47 | 33 | 6.2 | 4.3 | 9.7 | 0.0 |
| Classroom 2 | 437 | 10.7 | 23 | 16 | 5.8 | 3.6 | 8.1 | 0.0 |
| Classroom 3 | 860 | 9.8 | 38 | 27 | 1.0 | 3.4 | 7.5 | 4.5 |
| Classroom 3* | 860 | 9.8 | 38 | 27 | 1.0 | 3.4 | 7.5 | 0.0 |
| Classroom 4 | 901 | 10.7 | 53 | 37 | 3.7 | 4.0 | 8.9 | 0.0 |
| Conference Room | 437 | 10.7 | 16 | 11 | 3.9 | 2.7 | 7.3 | 0.0 |
| Lecture Hall 1 | 1818 | 10.0 | 120 | 60 | 4.3 | 3.3 | 9.7 | 0.0 |
| Lecture Hall 2 | 1861 | 13.1 | 128 | 64 | 6.9 | 2.6 | 7.7 | 0.0 |
| Office | 910 | 10.7 | 15 | 8 | 3.1 | 1.6 | 6.0 | 0.0 |
| Small Office 1 | 215 | 10.7 | 4 | 2 | 5.2 | 1.7 | 6.6 | 0.0 |
| Small Office 2 | 215 | 10.7 | 4 | 4 | 5.2 | 1.7 | 6.6 | 0.0 |

#### Classroom 1

Classroom 1 is a modestly-sized classroom used for small lectures or recitations. The room is served by a dedicated outdoor-air system with supplemental heating and cooling provided by in-room radiators and fan-coil units. Seating is free-standing chairs with attached writing boards, arranged in a grid and facing a chalkboard in the front of the room. The room has one exterior-facing wall with sealed windows.

Timeseries data is plotted in Figure 1, while daily energy and transmission-rate metrics are shown in 2.

#### Classroom 2

Classroom 2 is a small-sized classroom used for small lectures or recitations. The room is served by a dedicated outdoor-air system with supplemental heating and cooling provided by in-room radiators and fan-coil units. Seating is free-standing chairs with attached writing boards, arranged in a grid and facing a chalkboard in the front of the room. The room has one exterior-facing wall with sealed windows.

Timeseries data is plotted in Figure 3, while daily energy and transmission-rate metrics are shown in 4.

#### Classroom 3

Classroom 3 is a modestly sized classroom used for small lectures or recitations. The room is naturally ventilated (i.e., does not have a mechanically-provided source of outdoor air) with supplemental heating and cooling provided by in-room radiators and fan-coil units. In the back of the room, a free-standing HEPA air filter is installed, which can provided 4.5 ACH of airflow (as calculated from the nominal capacity of the unit and the room volume). For the results listed under "Classroom 3", we assume that this filter is active during nominally occupied hours (8am to 10pm each day). Seating is in free-standing chairs and tables arranged in rows facing a chalkboard in the front of the room. The room has one exterior-facing wall with operable windows.

Timeseries data is plotted in Figure 5, while daily energy and transmission-rate metrics are shown in 6.

#### Classroom 3*

Classroom 3* is the same room as "Classroom 3", except that we assume that the in-room HEPA air filter is *never* active. During the monitoring period, it is likely that the filter was active some but not all of the time, and thus the true transmission risk lies somewhere between the "Classroom 3" and "Classroom 3*" extremes.

Timeseries data is plotted in Figure 7, while daily energy and transmission-rate metrics are shown in 8.

#### Classroom 4

Classroom 4 is a large-sized classroom used for small lectures or recitations. The room is served by a dedicated outdoor-air system with supplemental heating and cooling provided by in-room radiators and fan-coil units. Seating is free-standing chairs with attached writing boards, arranged in a grid and facing a chalkboard in the front of the room. The room has one long exterior-facing wall with sealed windows.

Timeseries data is plotted in Figure 9, while daily energy and transmission-rate metrics are shown in 10.

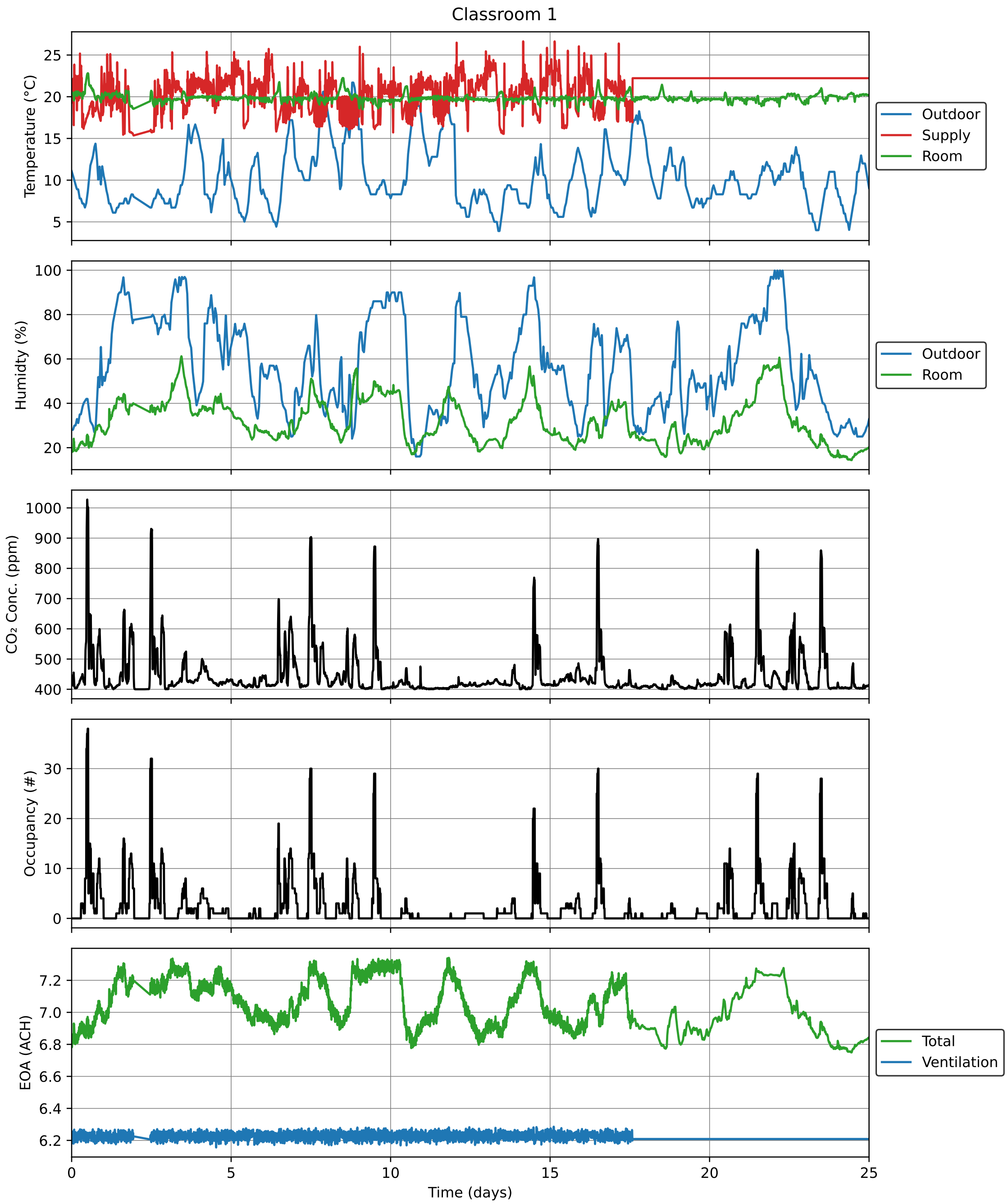

Figure 1: Timeseries data for Classroom 1 over the study period.

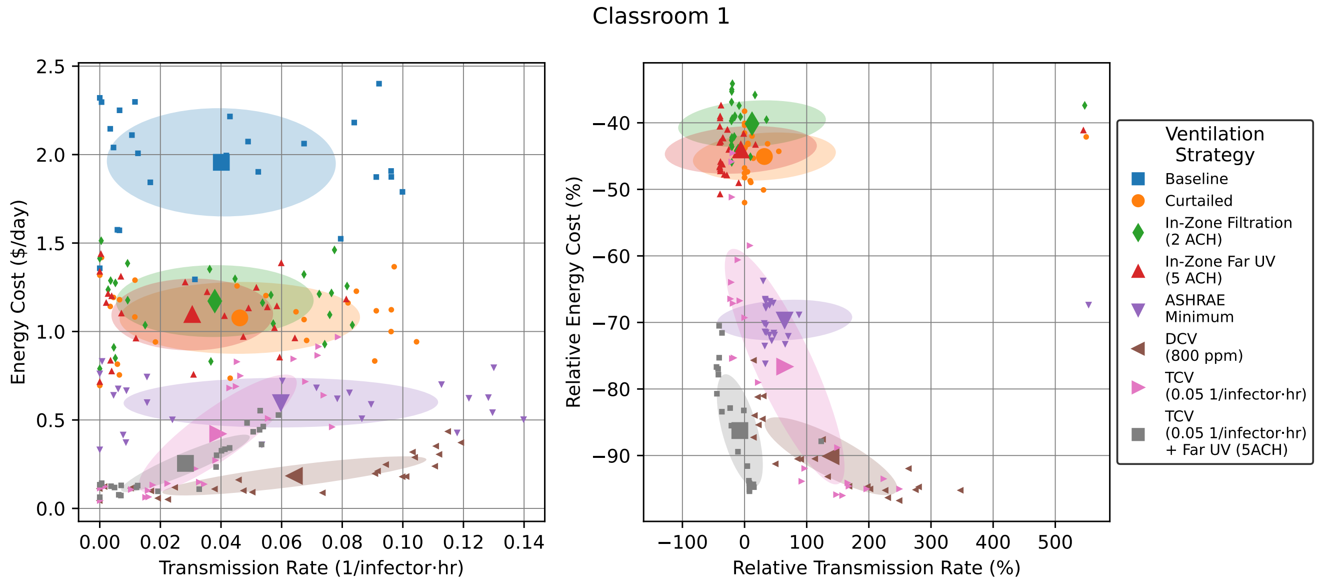

Figure 2: Energy versus transmission rate tradeoffs for hypothetical ventilation scenarios in Classroom 1. Small points show individual daily values, while large points with error bars show mean ± standard deviation. Right plot is relative to "Baseline" for each day.

#### Conference Room

Conference Room is a small-sized classroom that has been set up to facilitate conferences or other small meetings. The room is served by a dedicated outdoor-air system with supplemental heating and cooling provided by in-room radiators and fan-coil units. Seating is in free-standing chairs arranged around a large oval-shaped conference room, with additional chairs along the back wall of the room. The room has one exterior-facing wall with sealed windows.

Timeseries data is plotted in Figure 11, while daily energy and transmission-rate metrics are shown in 12.

#### Lecture Hall 1

Lecture Hall 1 is a moderately sized lecture hall used for larger lectures. Ventilation and cooling is provided by an air-handling unit (AHU), which is set to 100% outdoor-air operation throughout the monitoring period. Heating is provided by radiators. Seating is in attached seats with stowable writing boards arranged in slightly curved rows facing a chalkboard at the front of the room. There is slight vertical separation between the rows to facilitate view. The room has two exterior-facing walls with operable windows.

Timeseries data is plotted in Figure 13, while daily energy and transmission-rate metrics are shown in 14.

#### Lecture Hall 2

Lecture Hall 2 is a moderately sized lecture hall used for larger lectures. Ventilation and cooling is provided by an air-handling unit (AHU), which is set to 100% outdoor-air operation throughout the monitoring period. Heating is provided by radiators. Seating is in attached seats with stowable writing boards arranged in slightly curved rows facing a chalkboard at the front of the room. There is modest

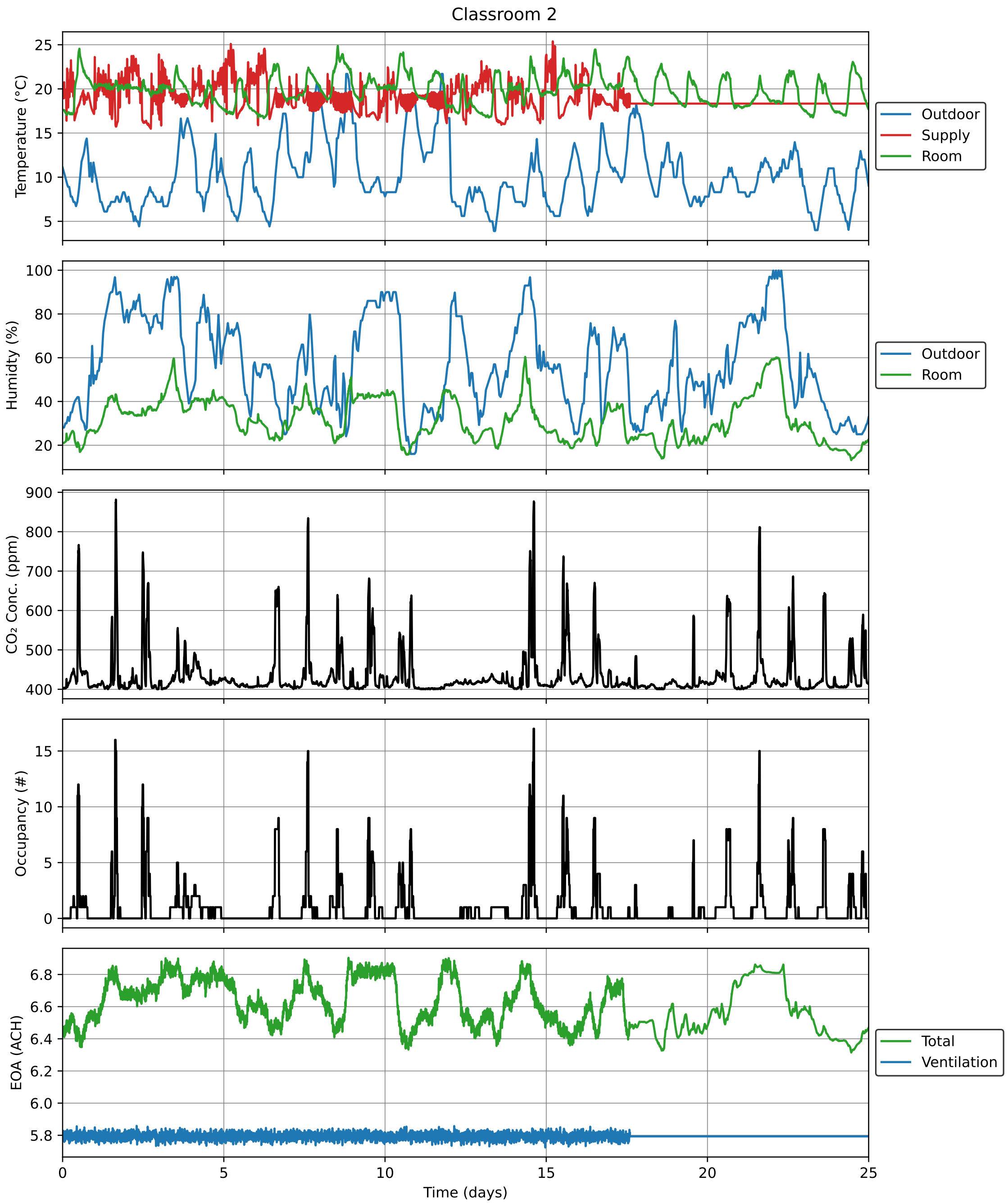

Figure 3: Timeseries data for Classroom 2 over the study period.

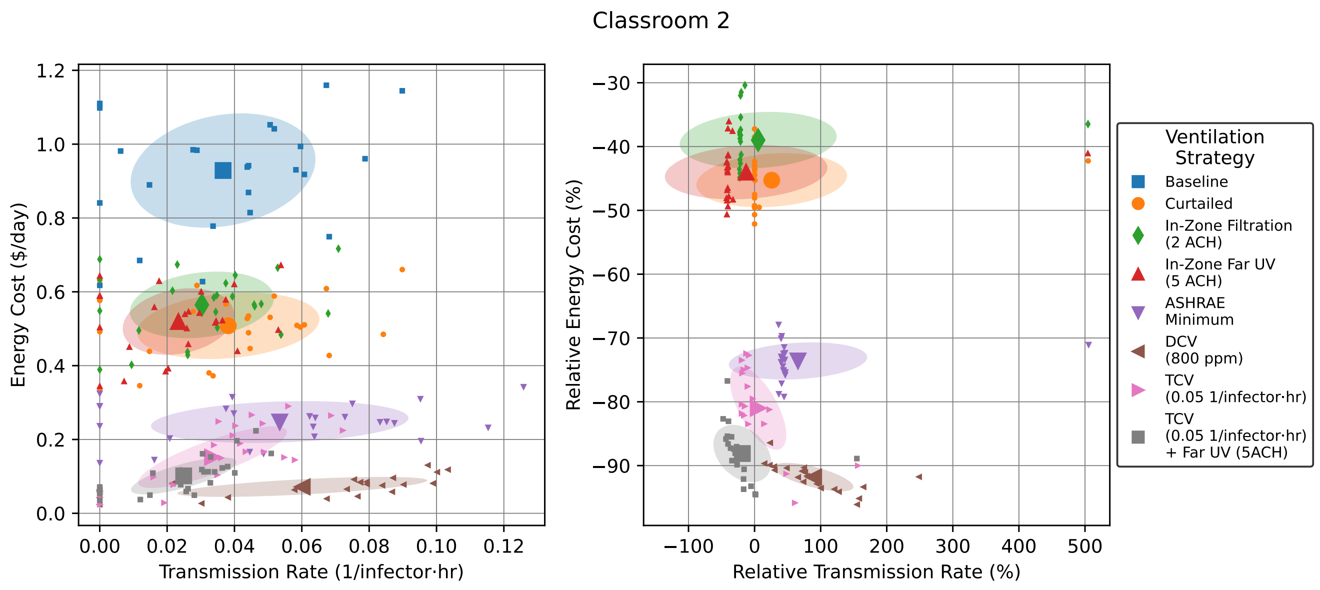

Figure 4: Energy versus transmission rate tradeoffs for hypothetical ventilation scenarios in Classroom 2. Small points show individual daily values, while large points with error bars show mean ± standard deviation. Right plot is relative to "Baseline" for each day.

vertical separation between the rows to facilitate view. The room has three exterior-facing walls with operable windows.

Timeseries data is plotted in Figure 15, while daily energy and transmission-rate metrics are shown in 16.

#### Office

Office is a modestly sized shared office used by graduate students. As a result of remodeling, the room is served by two separate dedicated outdoor-air systems. (Unfortunately, only one of the systems has a flow measurement, and so we assume that ventilation measurements are *not* available for this room.) Similarly, cooling is provided by two in-room fan-coil units which control to different setpoints (thus creating a measurable temperature gradient within the room). Heating is via radiators. Seating is primarily in office chairs at desks arranged in rows separated by cubicle walls. Each row has space for one or two occupants on either side. There is also a small kitchen area with a table, sink, microwave, and coffee maker in the middle of the room. There is one long exterior-facing wall with sealed windows.

Timeseries data is plotted in Figure 17, while daily energy and transmission-rate metrics are shown in 18.

#### Small Office 1

Small Office 1 is a small office space and waiting area near additional small offices. The room is served by a dedicated outdoor-air system with supplemental heating and cooling provided by in-room radiators and fan-coil units. Seating is at one chair behind a desk and three additional chairs on the other side of the room for waiting occupants. The room has one exterior-facing wall with sealed windows.

Timeseries data is plotted in Figure 19, while daily energy and transmission-rate metrics are shown in 20.

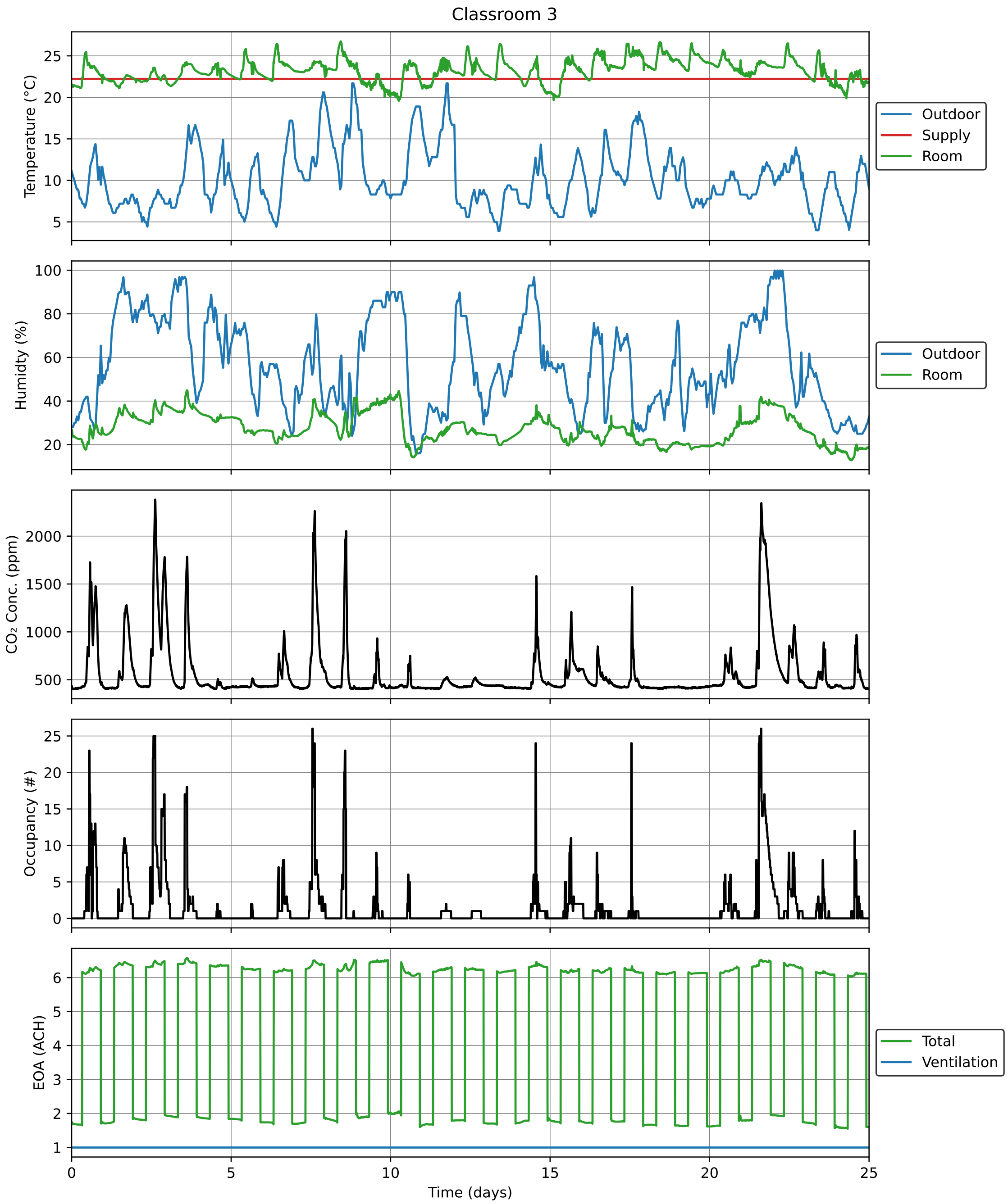

Figure 5: Timeseries data for Classroom 3 over the study period.

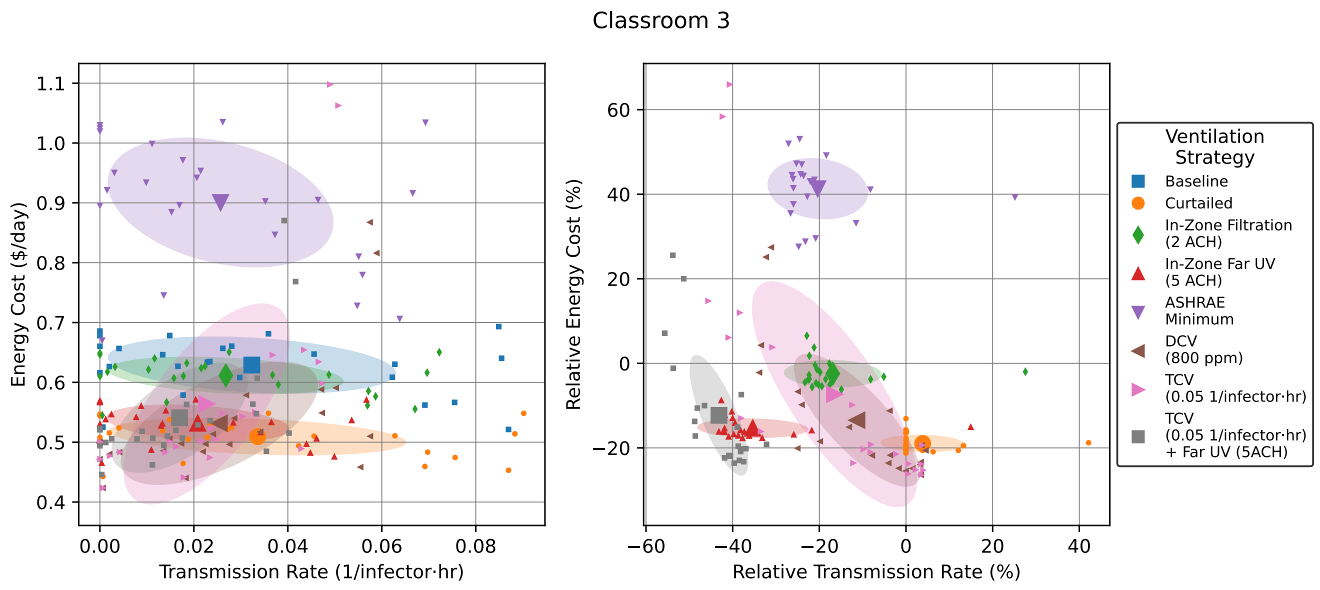

Figure 6: Energy versus transmission rate tradeoffs for hypothetical ventilation scenarios in Classroom 3. Small points show individual daily values, while large points with error bars show mean ± standard deviation. Right plot is relative to "Baseline" for each day.

##### Small Office 2

Small Office 2 is a small office space and meeting area. The room is served by a dedicated outdoor-air system with supplemental heating and cooling provided by in-room radiators and fan-coil units. Seating is at one chair behind a desk and at four additional chairs arranged around a small conference table. The room has one exterior-facing wall with sealed windows.

Timeseries data is plotted in Figure 21, while daily energy and transmission-rate metrics are shown in 22.

#### 2 Transmission Modeling Details

For airborne transmission, the production of pathogen, $P(r)$, comes from the exhaled breath of infectious individuals. This is given as

$$P(r) = Q_b n_d(r) V_d(r) p_m(r) c_v(r), \tag{1}$$

where $Q_b$ is the breathing flow rate of the individuals; $n_d(r)$ is the number density of pathogens per volume of breath, which is known to vary with factors that include respiratory activity, time since infection, etc.; $V_d(r)$ is the volume of the aerosol droplets; $p_m$ is a mask penetration factor which accounts for the proportion of pathogen that may be filtered out by the mask (where a value of 1 means all pathogen escapes the mask and 0 means all pathogen is filtered by the mask) [2]; and $c_v(r)$ is the pathogen concentration in the droplets.

According to this model, the steady-state value of pathogen concentration if one infector is present is

$$C_s(r) = \frac{P(r)}{\lambda_{\text{EOA}}(r) V}, \tag{2}$$

where $\lambda_{\text{EOA}}$ was defined in the main text.

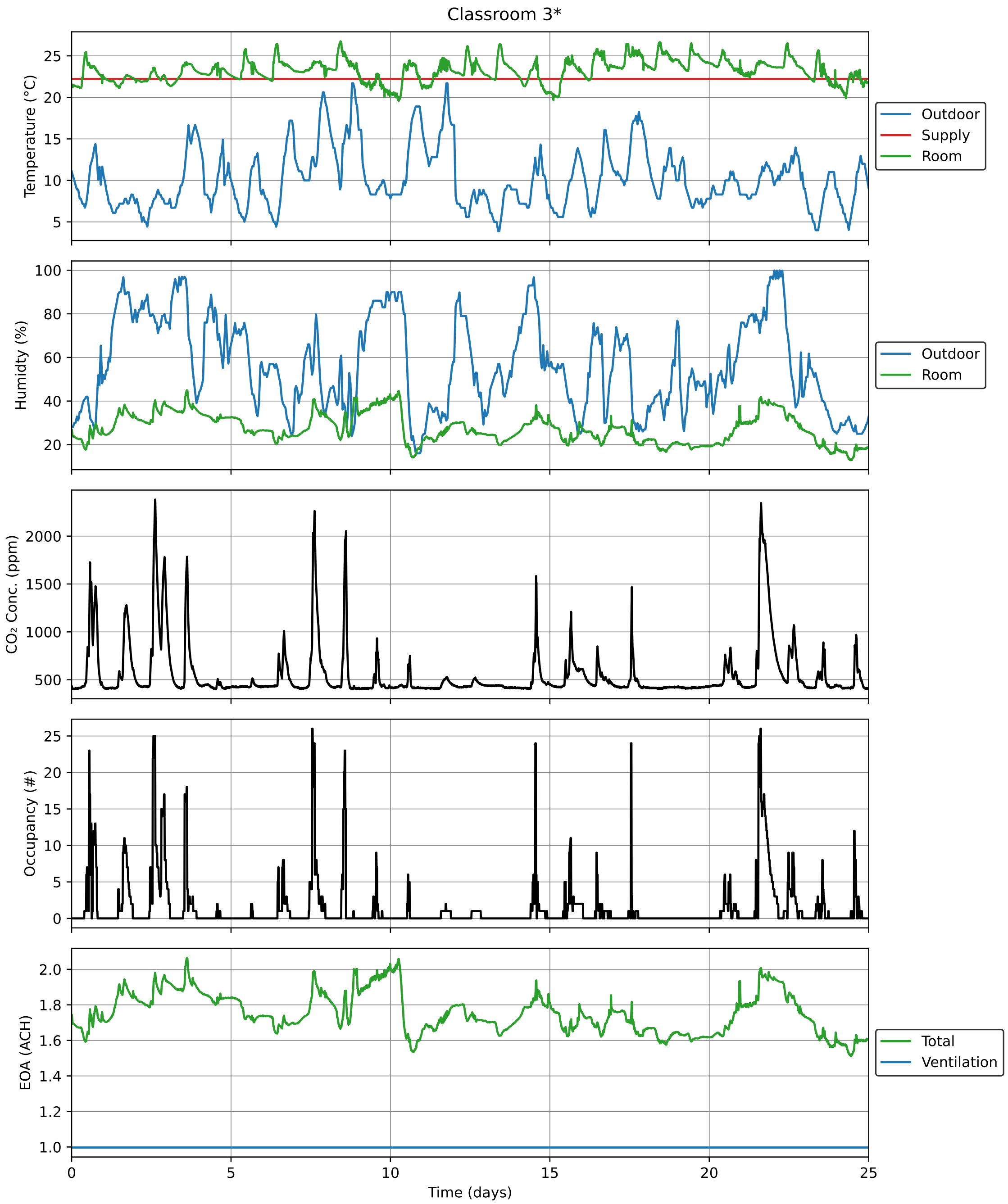

Figure 7: Timeseries data for Classroom 3* over the study period.

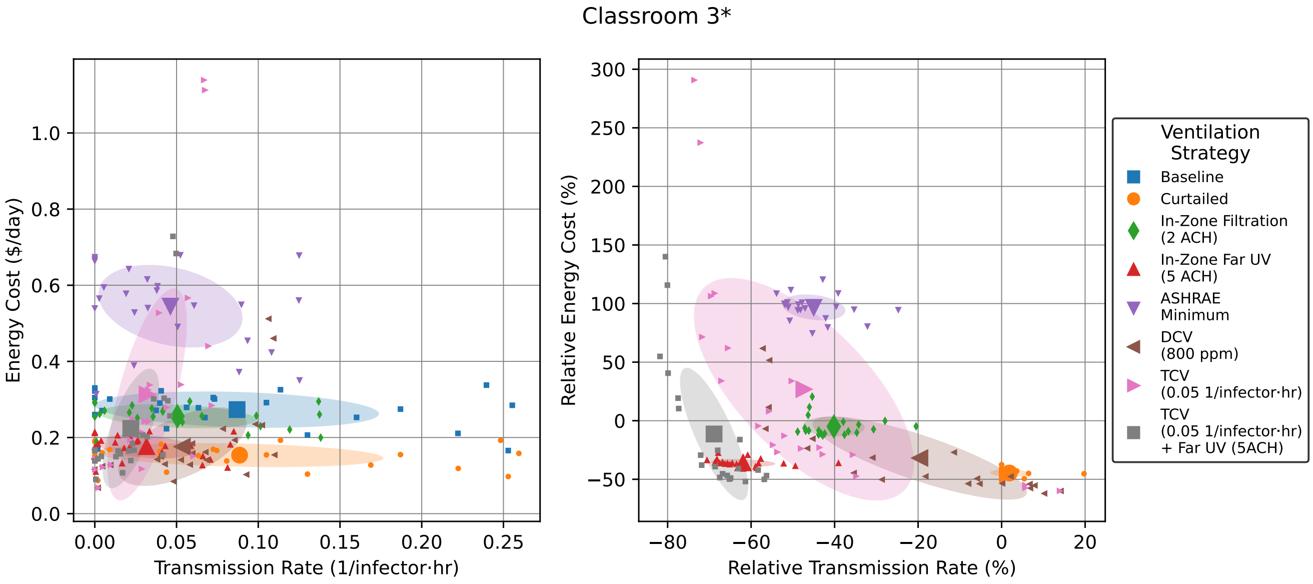

Figure 8: Energy versus transmission rate tradeoffs for hypothetical ventilation scenarios in Classroom 3*. Small points show individual daily values, while large points with error bars show mean ± standard deviation. Right plot is relative to "Baseline" for each day.

We note that the distribution of a ventilation system (location of inflow and outflow) may impact the risk of spreading a contaminant. This risk increase is more complicated and must be analyzed by solving for the airflow patterns using computational fluid dynamics (CFD) techniques. If a ventilation system is not laid out well, this can lead to an increase of transmission risk upon an increase in ventilation. Alternatively, ventilation strategies like displacement ventilation deliberately cause stratification of the air within a room, which can lead to further complications. These factors are important to consider when designing and analyzing transmission in indoor spaces. However, all spaces monitored in this study make use of mixing ventilation, which is intentionally designed to achieve thorough mixing. The well-mixed assumption has been shown to capture the dominant transmission dynamics and leads to simple guidelines, which is why we focus on it in this study.

### 3 Safety Guideline Derivation

The safety guideline derivation is based on [3] and a more detailed derivation of the mathematics, which is also presented here, is given in [4].

The safety guideline is based on a linear approximation to the exponential dose-response model, which is derived here. The infection process has two main steps. The first step involves a person being exposed to pathogens. This step is mainly dictated by the physics of aerosol transport, which we have good models to quantify. The second step involves the pathogen evading the body's defenses and actually causing an infection. This step is mainly dictated by pathogen and human biology, which is harder to quantify is thus often lumped into parameters that can be fit to epidemiology data.

To quantify exposure, we will assume that all people and pathogens act independently. We also assume that the pathogen is randomly distributed in a room. Therefore, the number of pathogens that a person breathes in during an event is Poisson-distributed. Therefore, the probability that $v$ pathogens

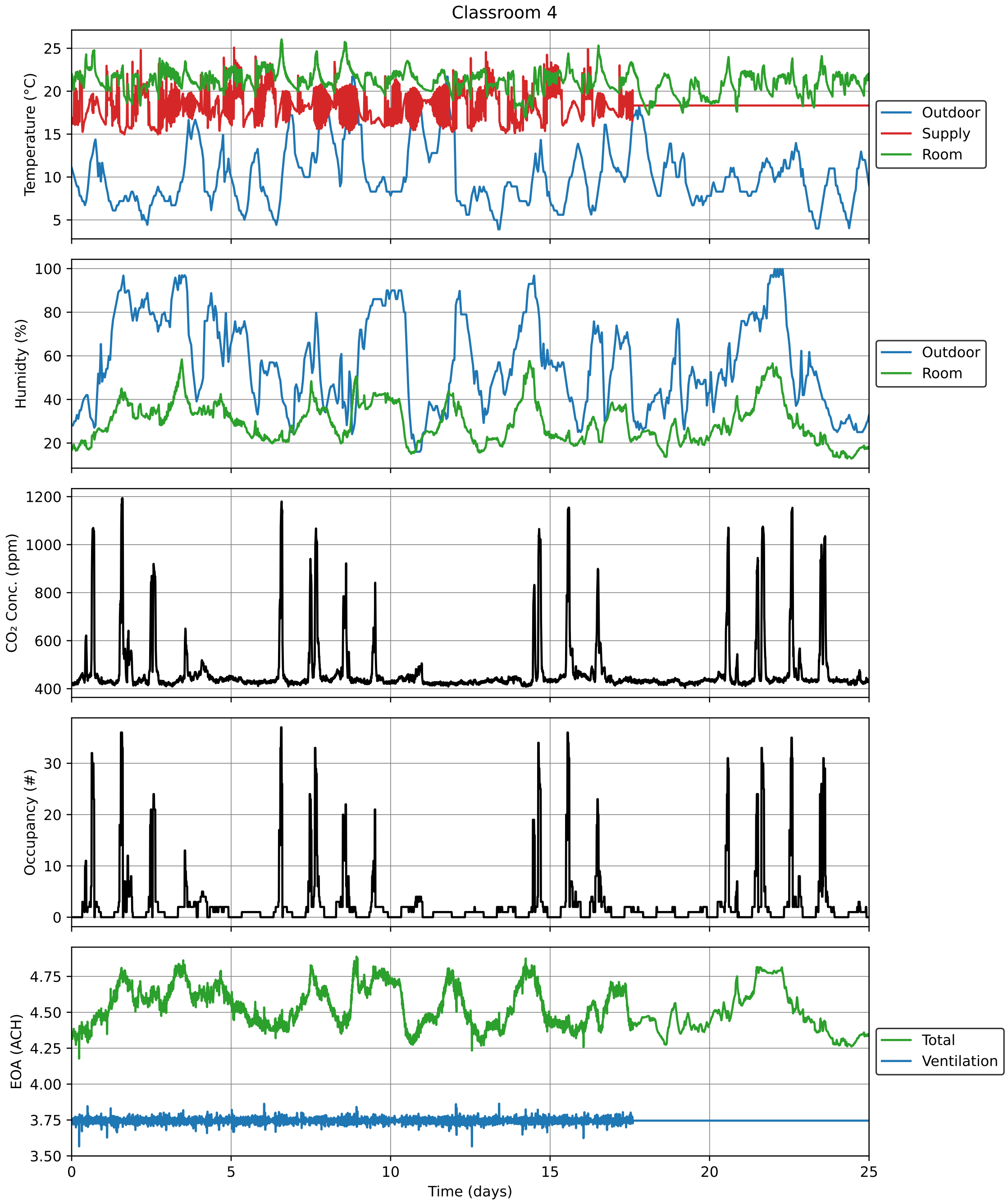

Figure 9: Timeseries data for Classroom 4 over the study period.

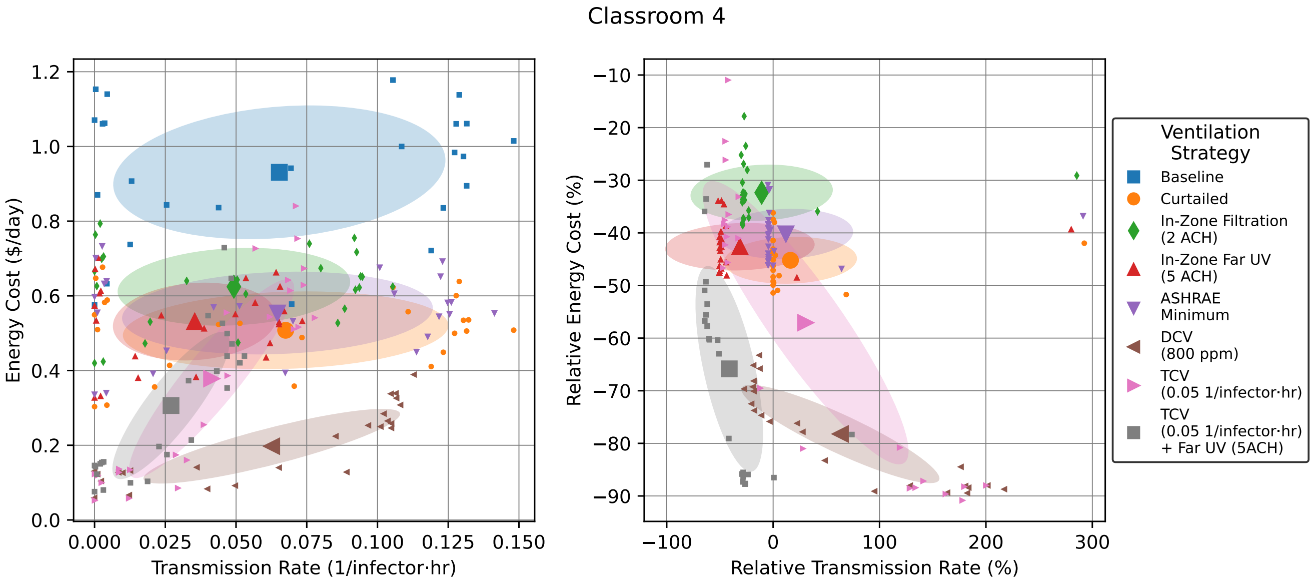

Figure 10: Energy versus transmission rate tradeoffs for hypothetical ventilation scenarios in Classroom 4. Small points show individual daily values, while large points with error bars show mean ± standard deviation. Right plot is relative to "Baseline" for each day.

are inhaled is given by

$$P_{inhale}(v) = \frac{\lambda^v e^{-\lambda}}{v!}, \tag{3}$$

where $\lambda$ is the expected value of pathogen consumption which is called the average dose in the dose-response literature. The average dose is equal to the product of the amount of air breathed in and the concentration of the pathogen, $C(r, t)$. We define the amount of air a person is breathing in to be $Q_b$, which we assume to be constant in time. If we include a factor to account for potential mask usage, whose filtration efficiency may be size-dependent, the concentration of pathogen is given by $\int_0^\tau \int_0^\infty C(r, t) dr dt$. Therefore, the average dose is

$$\lambda = Q_b \int_0^\tau \int_0^\infty C(r, t) dr dt. \tag{4}$$

To quantify infection, we will assume that all exposures have the same probability that each consumed pathogen will initiate an infection, but this probability may be size-dependent. Therefore, the number of pathogens that cause an infection is binomial-distributed. Therefore, the probability that there will be $f$ infections from the $v$ pathogens inhaled is

$$P_{infect}(f, v) = \frac{v!}{f!(v-f)!} c_i^f (1 - c_i)^{v-f}, \tag{5}$$

where $c_i$ is the probability that a pathogen causes an infection.

We can then multiply these probabilities and sum over all values of $f$ and $v$ that would cause an infection in order to get the total probability of infection:

$$P_{infection} = \sum_{f=f_{min}}^{\infty} \sum_{v=f}^{\infty} P_{inhale}(v) P_{infect}(f, v) \tag{6}$$

$$= \sum_{f=f_{min}}^{\infty} \sum_{v=f}^{\infty} \left( \frac{\lambda^v e^{-\lambda}}{v!} \right) \left( \frac{v!}{f!(v-f)!} c_i^f (1 - c_i)^{v-f} \right), \tag{7}$$

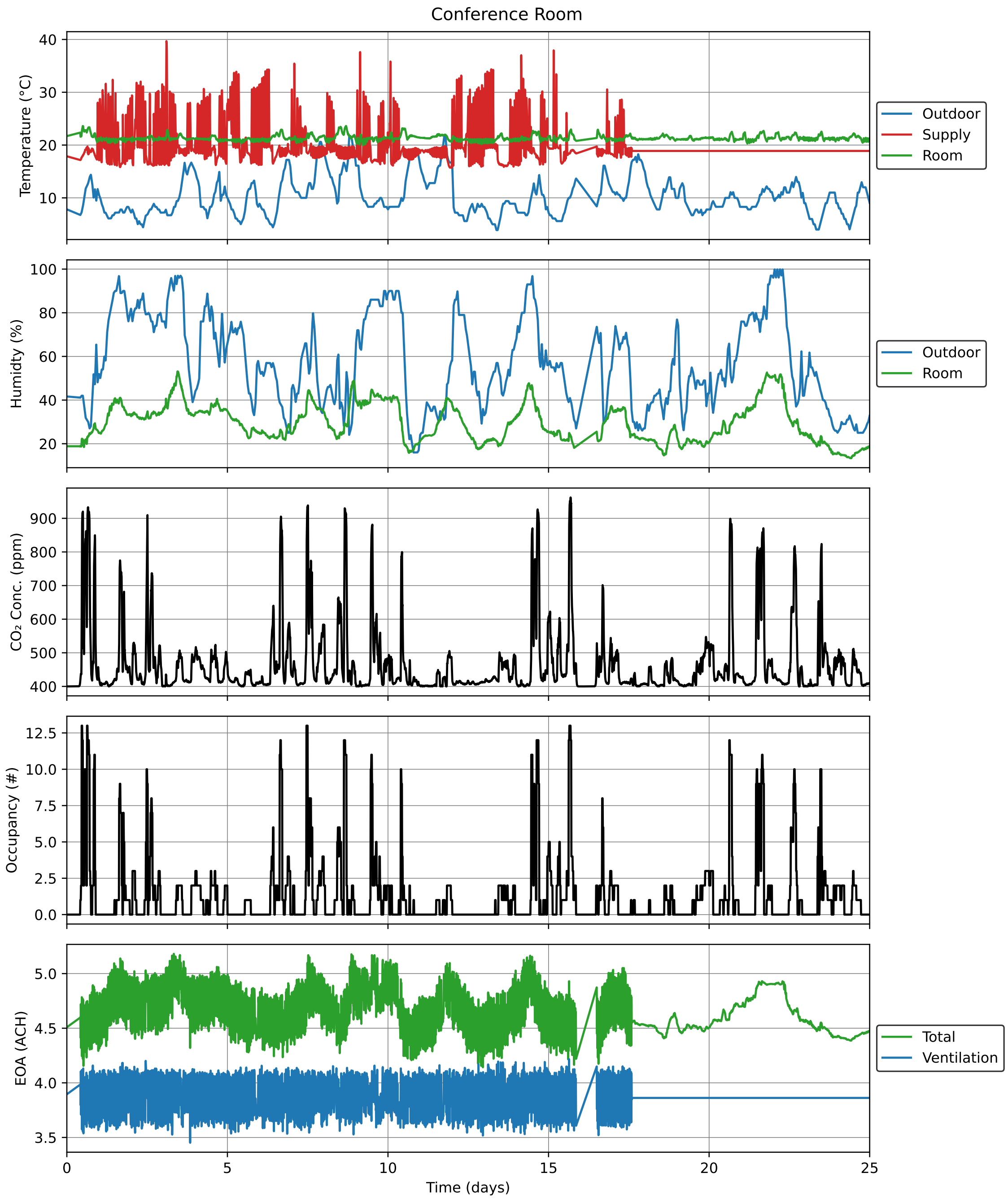

Figure 11: Timeseries data for Conference Room over the study period.

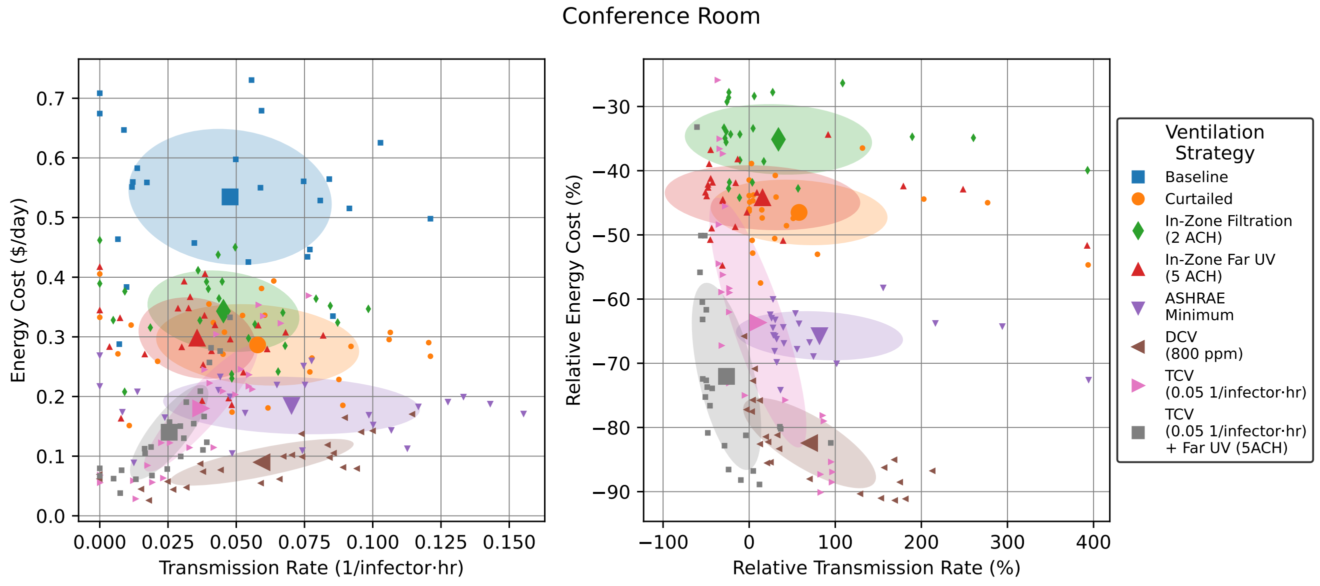

Figure 12: Energy versus transmission rate tradeoffs for hypothetical ventilation scenarios in Conference Room. Small points show individual daily values, while large points with error bars show mean ± standard deviation. Right plot is relative to "Baseline" for each day.

where $f_{min}$ is the minimum number of infections required to cause the illness or disease associated with the infection.

We can simplify this by cleverly rearranging the right hand side, as done in [4], to be

$$P_{infection} = \sum_{f=f_{min}}^{\infty} \frac{(\lambda c_i)^f e^{-\lambda c_i}}{f!} \sum_{v=f}^{\infty} \frac{(\lambda(1-c_i))^{v-f} e^{-\lambda(1-c_i)}}{(v-f)!}. \tag{8}$$

To evaluate the rightmost sum we can pull out the $e^{-\lambda(1-c_i)}$ and begin to expand it as

$$\sum_{v=f}^{\infty} \frac{(\lambda(1-c_i))^{v-f} e^{-\lambda(1-r)}}{(v-f)!} = e^{-\lambda(1-c_i)} \left(1 + (\lambda(1-c_i)) + \frac{(\lambda(1-c_i))^2}{2!} + \frac{(\lambda(1-c_i))^3}{3!} + \ldots\right). \tag{9}$$

Recognizing the sum as the exponential Taylor series we get

$$\sum_{v=f}^{\infty} \frac{(\lambda(1-c_i))^{v-f} e^{-\lambda(1-c_i)}}{(v-f)!} = e^{-\lambda(1-c_i)} e^{\lambda(1-c_i)} = 1. \tag{10}$$

Therefore, if we also assume that one infection is enough to provoke the illness/disease, our probability of infection is now just

$$P_{infection} = \sum_{f=1}^{\infty} \frac{(\lambda c_i)^f e^{-\lambda c_i}}{f!}. \tag{11}$$

We notice that this is just the sum of the probability mass function of a Poisson distribution, just missing the $f = 0$ term. Therefore, since the sum of the probability must equal one, this is equal to one minus the $f = 0$ term:

$$P_{infection} = 1 - e^{\lambda c_i}. \tag{12}$$

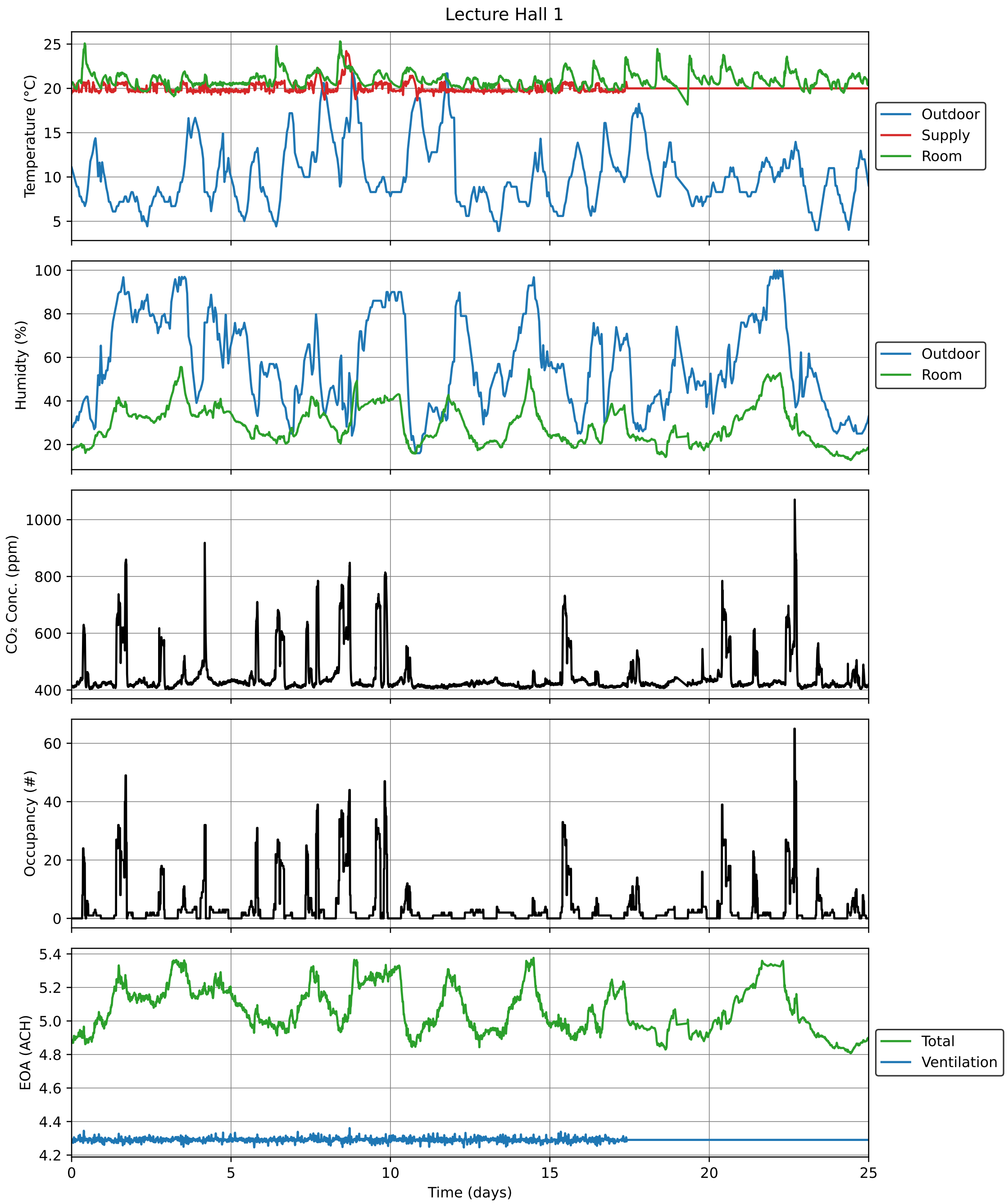

Figure 13: Timeseries data for Lecture Hall 1 over the study period.

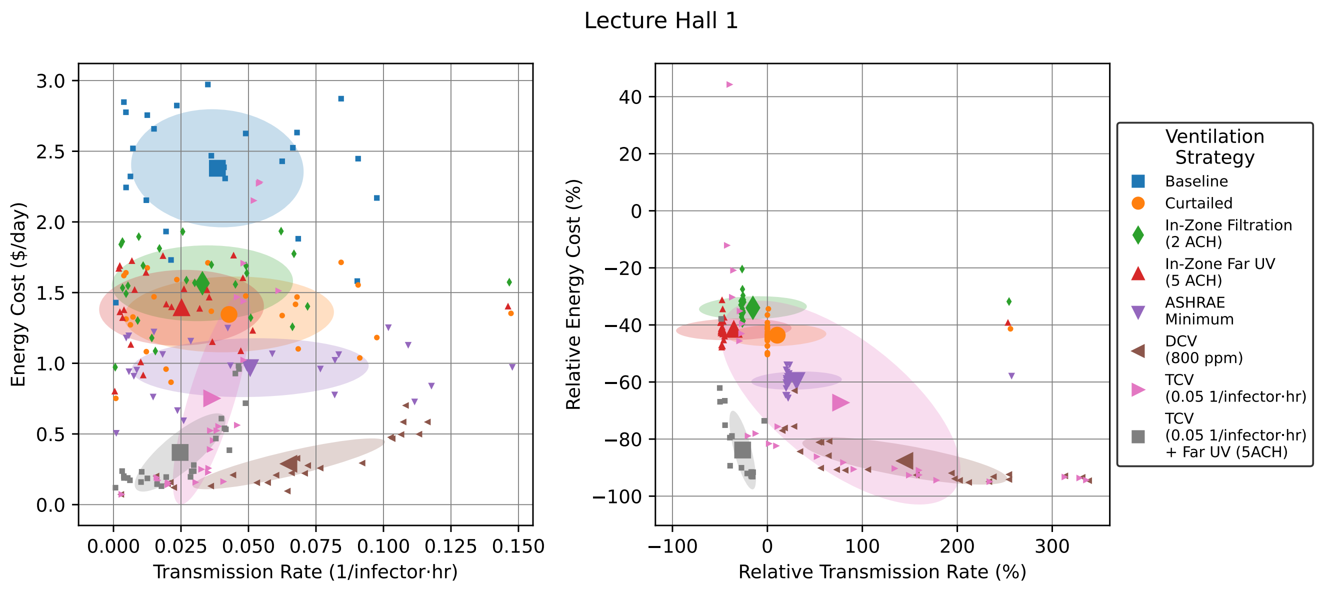

Figure 14: Energy versus transmission rate tradeoffs for hypothetical ventilation scenarios in Lecture Hall 1. Small points show individual daily values, while large points with error bars show mean ± standard deviation. Right plot is relative to "Baseline" for each day.

We now see how $c_i$ relates to the idea of "infection quanta" from the literature [5]. Since $\lambda$ represents amount pathogen, $c_i$ represents the infection quanta per pathogen. We also see that $c_i^{-1}$ is the amount of pathogen required to cause an infection with probability $1 - e^{-1} = 63\%$. Since $c_i$ may also be size-dependent, we can lump it in with the size integral to get

$$Q_b \int_0^\tau \int_0^\infty C(r,t) c_i dr dt. \tag{13}$$

To get a simple formula, we can now Taylor expand the exponential term in Equation (12) up to linear order to get

$$P_{infection} = Q_b \int_0^\tau \int_0^\infty C(r,t) c_i(r) dr dt. \tag{14}$$

We now add several terms to account for additional physical and biological effects. Since occupants may be wearing masks, the size-dependent mask penetration factor, $p_m(r)$, multiplies the pathogen concentration. Also, it has been observed that different subpopulations of people may be more susceptible to certain viruses and strains then others. To account for this, we also add a relative susceptibility, $s_r$, for rescaling the transmission rate.

After incorporating these effects, we now have

$$P_{infection} = Q_b s_r \int_0^\tau \int_0^\infty C(r,t) p_m(r) c_i(r) dr dt. \tag{15}$$

We then define the airborne transmission rate as

$$\beta_a(t) = Q_b s_r \int_0^\infty C(r,t) p_m(r) c_i(r) dr. \tag{16}$$

Thus, the expected number of transmission to a single person is

$$\int_0^\tau \beta_a(t) dt. \tag{17}$$

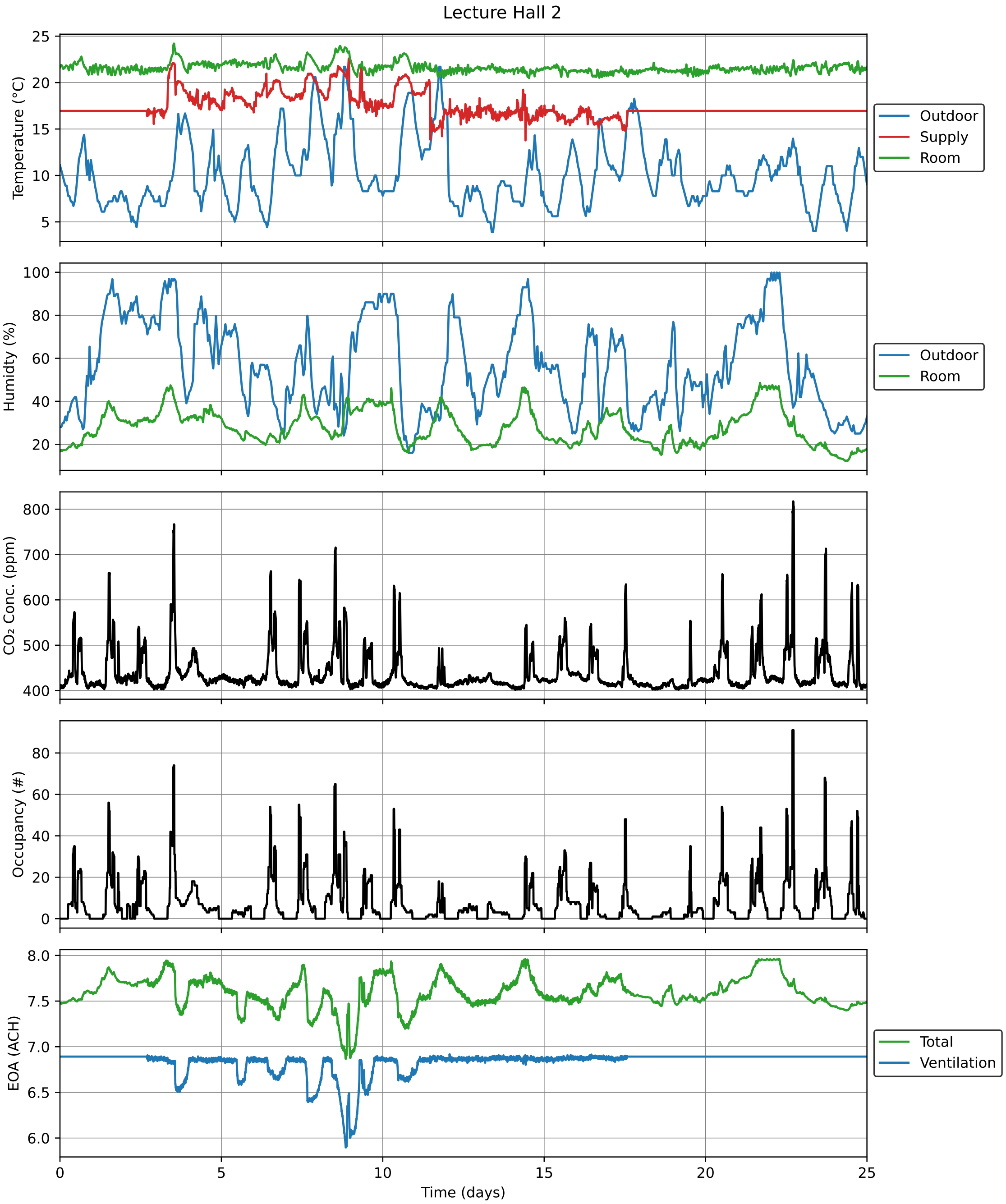

Figure 15: Timeseries data for Lecture Hall 2 over the study period.

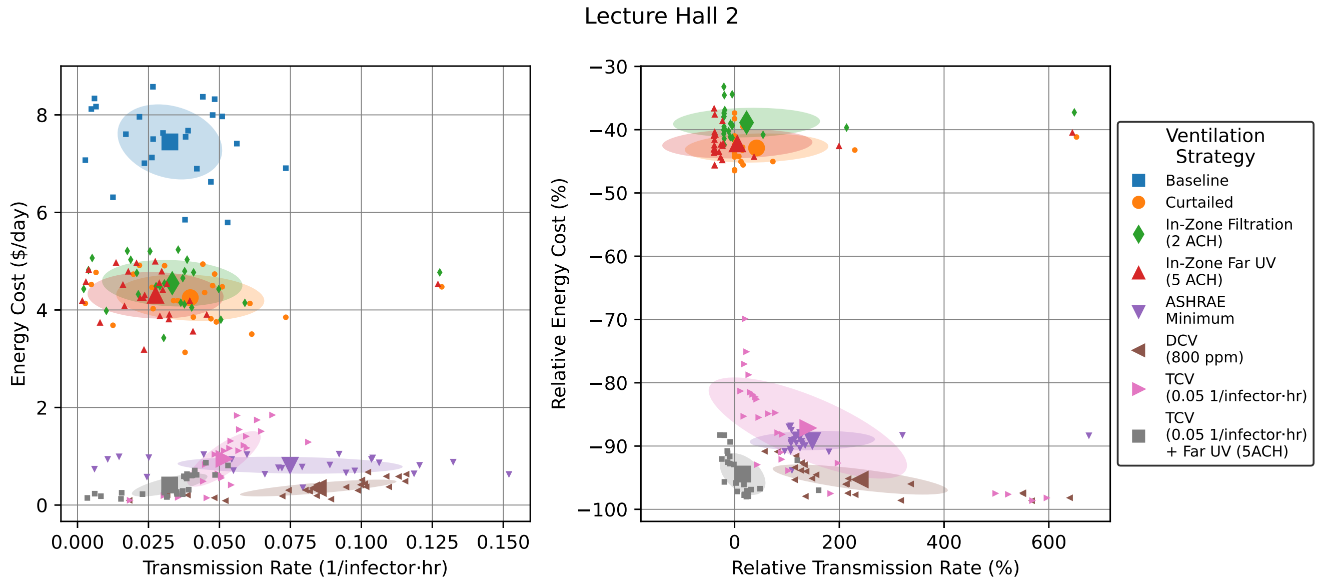

Figure 16: Energy versus transmission rate tradeoffs for hypothetical ventilation scenarios in Lecture Hall 2. Small points show individual daily values, while large points with error bars show mean ± standard deviation. Right plot is relative to "Baseline" for each day.

Since a single person can only be infected at most once (for the timescales that we are interested in) we treat an expected number of transmission of 1 and more than 1 on the same footing. Therefore, we can use this expected number of transmissions as an upper bound on the probability of transmission being at least 1 due to Markov's inequality.

We can now define an indoor reproductive number, $\mathcal{R}_{in}(\tau)$, as the expected number of transmissions in a room of $N_s$ susceptible people to be

$$\mathcal{R}_{in}(\tau) = N_s \int_0^\tau \beta_a(t)dt. \tag{18}$$

To set a specific safety guideline, we will typically want to bound $\mathcal{R}_{in}(\tau)$ by a maximum tolerance, $\epsilon$.

We can define a simple formula for the safety guideline using the steady-state pathogen concentration, which is valid when $\tau >> \lambda_{EOA}^{-1}$.

The steady-state approximate transmission rate now becomes

$$\bar{\beta}_a = Q_b s_r \int_0^\infty C_s(r) p_m(r) c_i(r) dr \tag{19}$$

$$= \frac{Q_b^2 s_r}{V} \int_0^\infty \frac{n_d(r) V_d(r) p_m^2(r) c_i(r) c_v(r)}{\lambda_{EOA}(r)} dr \tag{20}$$

We now assume $p_m$ is not size-dependent and define the concentration of infection quanta as $n_q(r) = n_d(r)V_d(r)c_v(r)c_i(r)$ we integrate over all $r$ to get the total infection quanta in exhaled air as $C_q = \int_0^r n_q(r)dr$. We can plus this into Equation (20) to get

$$\bar{\beta}_a = s_r \frac{Q_b^2 p_m^2}{V} \frac{C_q}{\lambda_{EOA}(\bar{r})}, \tag{21}$$

where the effective droplet radius $\bar{r}$ is defined such that Equation (20) and (21) are equal.

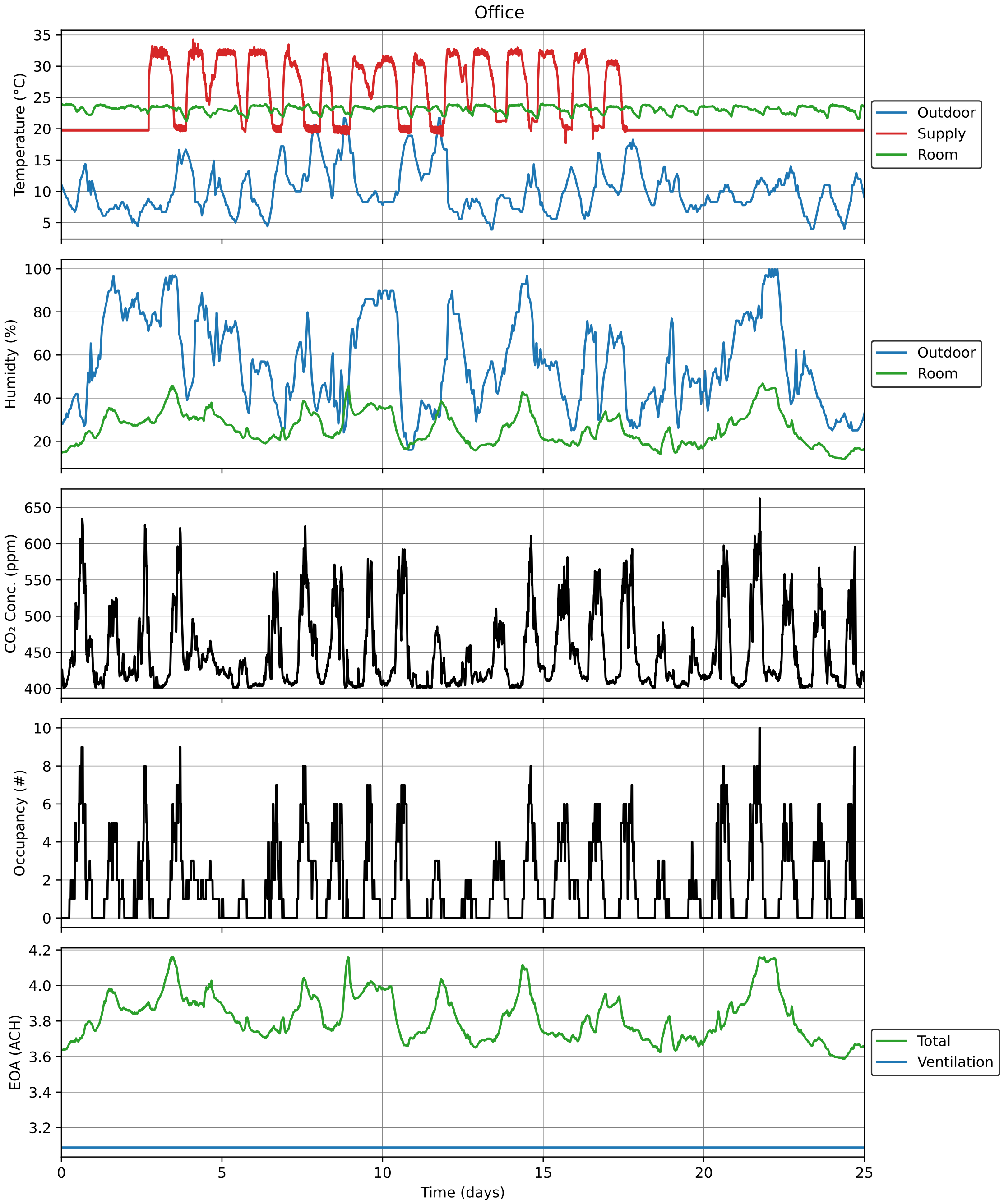

Figure 17: Timeseries data for Office over the study period.

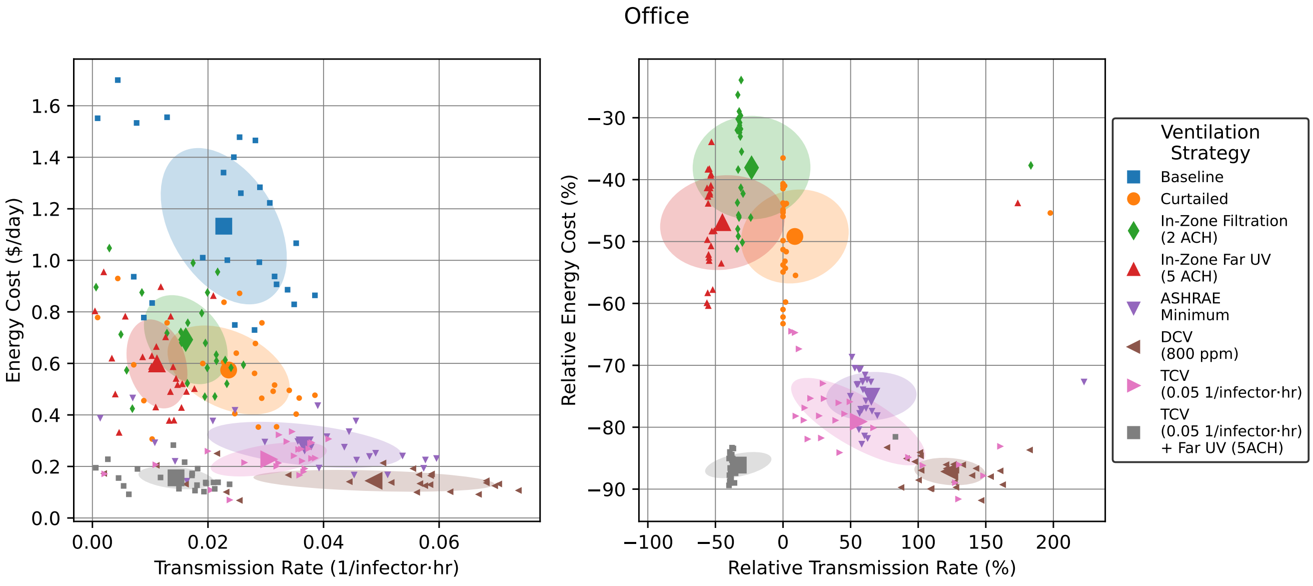

Figure 18: Energy versus transmission rate tradeoffs for hypothetical ventilation scenarios in Office. Small points show individual daily values, while large points with error bars show mean ± standard deviation. Right plot is relative to "Baseline" for each day.

We now arrive at our final safety guideline

$$N_s t < \epsilon \frac{\lambda_{EOA}(\bar{r}) V}{Q_b^2 p_m^2 C_q s_r}. \tag{22}$$

We note that all of the difficult to measure biological factors are lumped into a single parameter $C_q$ which can be fit to data for documented indoor spreading events and rescaled for different modes of respiration, as was first demonstrated for the COVID-19 wild type [6] and extended for the alpha, beta, gamma, delta, and omicron variants in our online app [7]. All other parameters are well-known. In addition, the parameters $\lambda_{EOA}$ and $p_m$ can be controlled to reduce transmissions.

### 4 Short-Range Transmission Risk

Specific values from the short-range transmission risk model presented in the main text that are used in the study are $Q_b = 0.56$ m$^3$/h, $V = 37.8$ m$^3$, and $\lambda_{\text{EOA}} = 6.6$ h$^{-1}$ (consisting of 5.6 h$^{-1}$ from ventilation and an assumed 1.1 h$^{-1}$ from deactivation and deposition), with measurements $\epsilon = 1.34$ at $x = 0.35$ m and $\epsilon = 1.7$ at $x = 1.1$ m. These values lead to least-squares estimate of $k = 3.00 \times 10^{-4}$ m.

As stated in the main text, the parameter $p_{\text{short}}$ represents the probability that a susceptible is directly within the short-range plume exhaled by each infector. This parameter is difficult to quantify, but as a conservative approximation, we make use of values reported by [8]. In [8], image-processing techniques were used to estimate the fraction of time that subway riders are oriented face-to-face, reporting fractions of 16.4% during rush hour and 52.8% during other times. Based on these values, we assume $p_{\text{short}} = 1$ for the small offices, 0.5 for the conference room, and 0.15 for all other spaces. Note that in all cases, we set $p_{\text{short}} = 0$ during times where there are fewer than two occupants (as no short-range transmission could take place). We assume in addition that $x = \min(x_0, \sqrt{0.8A/N_t})$ to reflect the fact that face-to-face contact would generally occur at distance $x_0 = 1$ m unless total occupant density $N_t/A$ is high enough that they must stand closer.

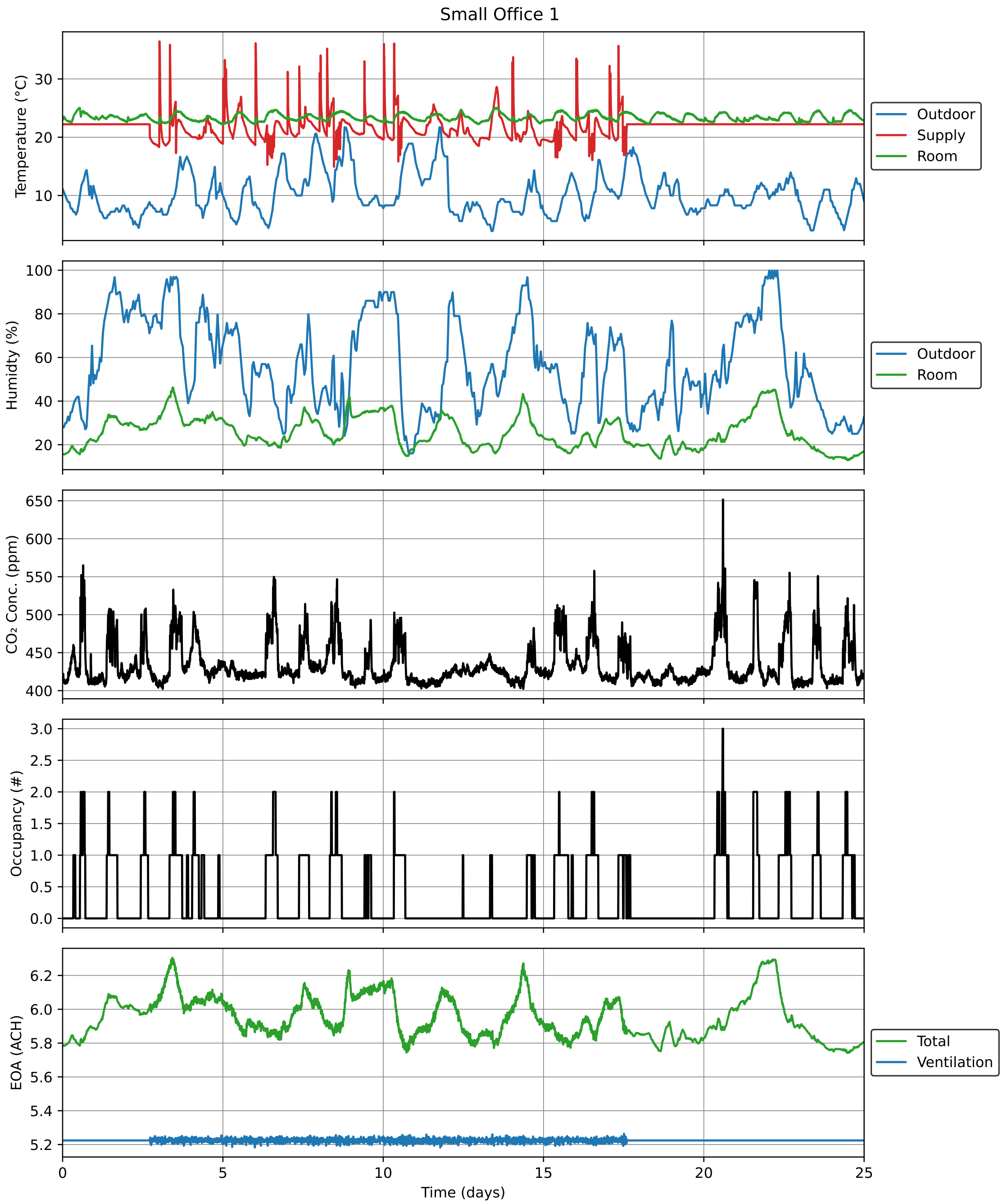

Figure 19: Timeseries data for Small Office 1 over the study period.

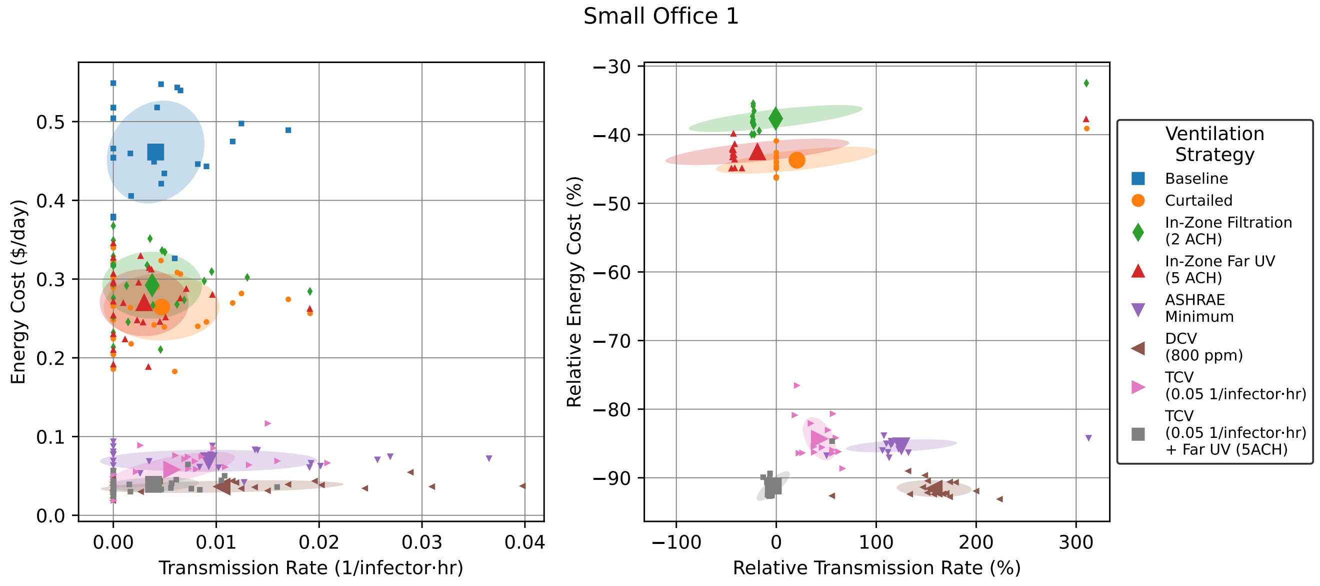

Figure 20: Energy versus transmission rate tradeoffs for hypothetical ventilation scenarios in Small Office 1. Small points show individual daily values, while large points with error bars show mean ± standard deviation.

Under the preceding assumptions, Figure 23 shows distributions for the short- and long-range transmission rates in each space. We see that across all spaces, the short-range transmission rates are always a small fraction of long-range transmission rates, thus justifying the focus on the latter route. The only possible outliers are the small offices, in which face-to-face contact is likely. Fortunately, this risk can be eliminated completely by requiring occupants to wear masks.

### 5 Hypothetical Ventilation Strategy Details

In this section, we provide additional details about the hypothetical operating scenarios. These include additional in-zone devices, as well as the demand-controlled and transmission-controlled ventilation scenarios, in which time-varying ventilation is simulated in accordance with a specific control objective.

#### 5.1 In-Zone Devices

For hypothetical simulation, we consider two types of in-zone disinfection devices: a free-standing HEPA air filter and upper-room far UV lamps. For the air filter, modeling is relatively straightforward. We assume that enough devices are installed to deliver the chosen volumetric airflow (set to 2 ACH in each room). We assume a removal efficiency of 0.999, which is multiplied by the nominal airflow to give EOA. Total power consumption is calculated assuming 0.65 W/CFM for the devices. Note that this scaling factor is in line with many residential air cleaners, which have power consumption between 0.1 and 2 W/CFM [10].

For far UV, the modeling is slightly more complicated. To calculate the equivalent outdoor air, we assume that sufficient lamps are installed to give a constant UV intensity throughout the volume of the room. This intensity then causes a constant decay rate for the microbes (commonly called the $k$ or $Z$ factor), which we assume to be equal to 5 $\mathrm{cm}^2/\mathrm{mJ}$ as measured by [11] for human coronaviruses. Thus, to deliver 5 ACH of EOA, the UV intensity of the room must be equal to 0.28 $\mu\mathrm{W/cm}^2$. This value is

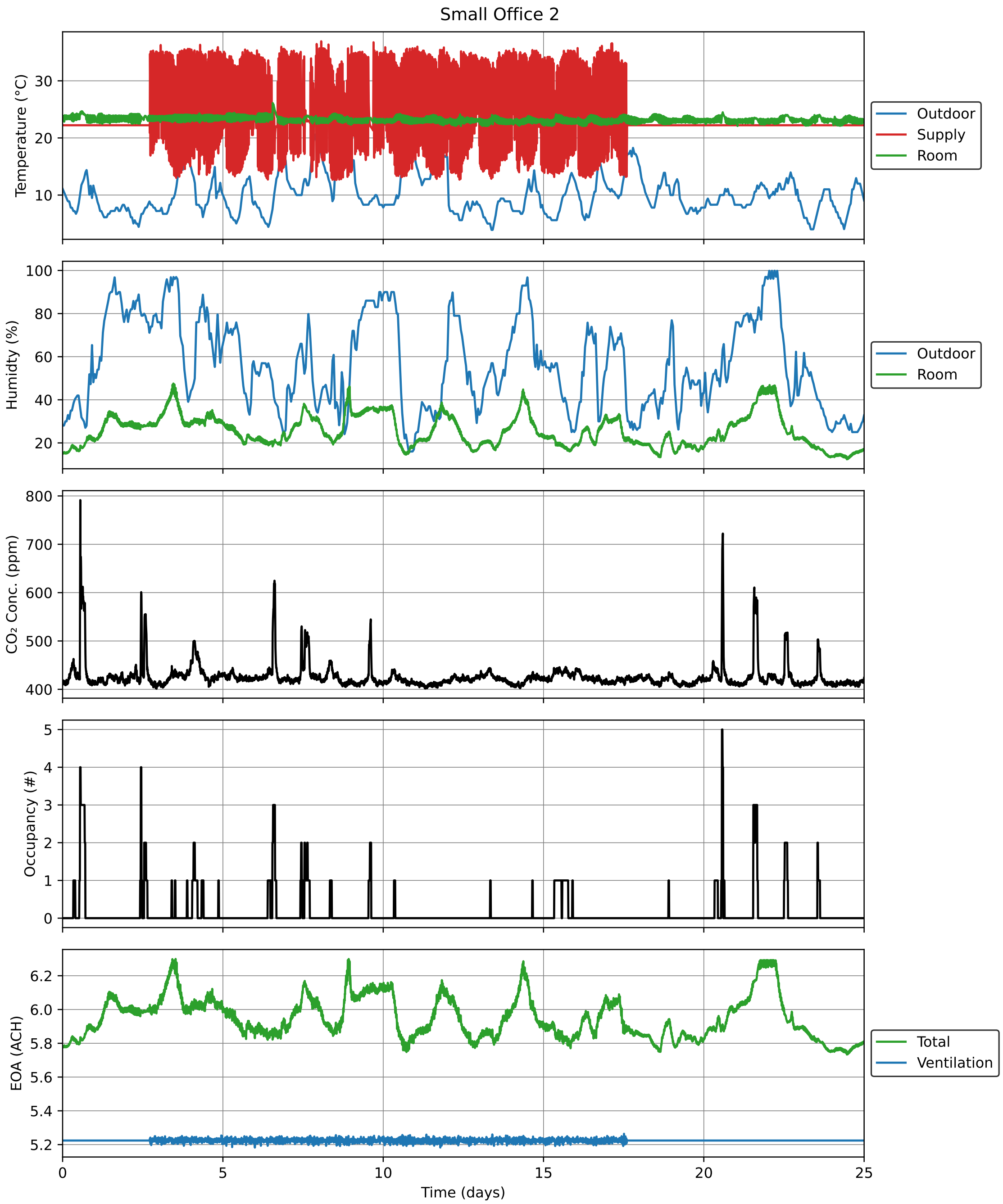

Figure 21: Timeseries data for Small Office 2 over the study period.

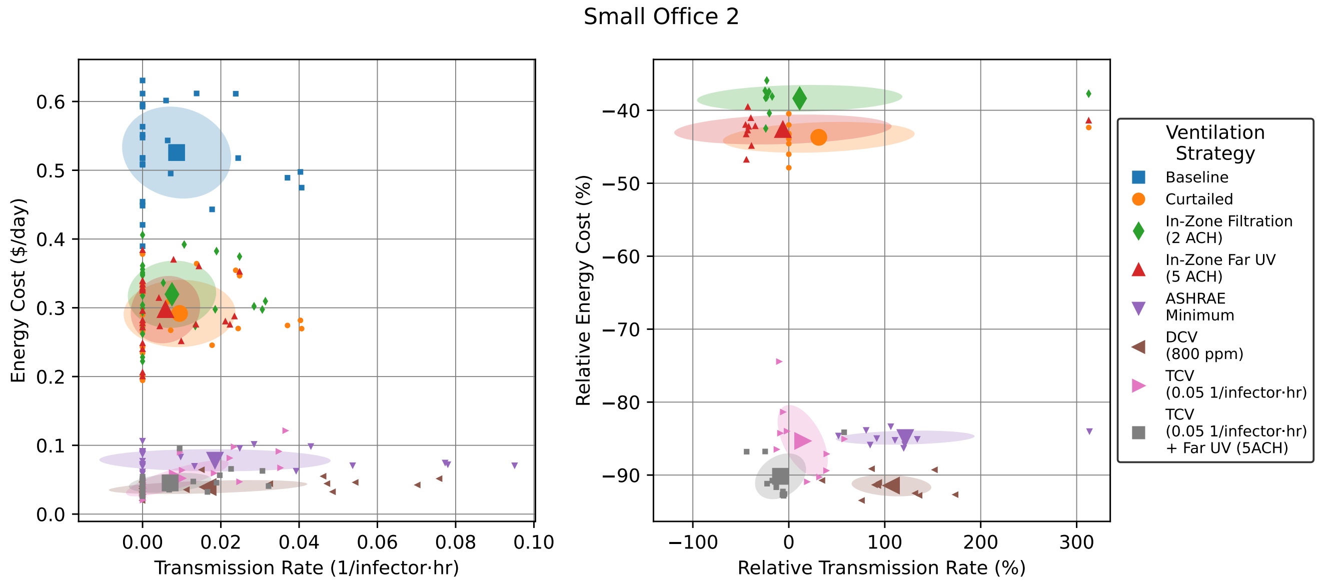

Figure 22: Energy versus transmission rate tradeoffs for hypothetical ventilation scenarios in Small Office 2. Small points show individual daily values, while large points with error bars show mean ± standard deviation. Right plot is relative to “Baseline” for each day.

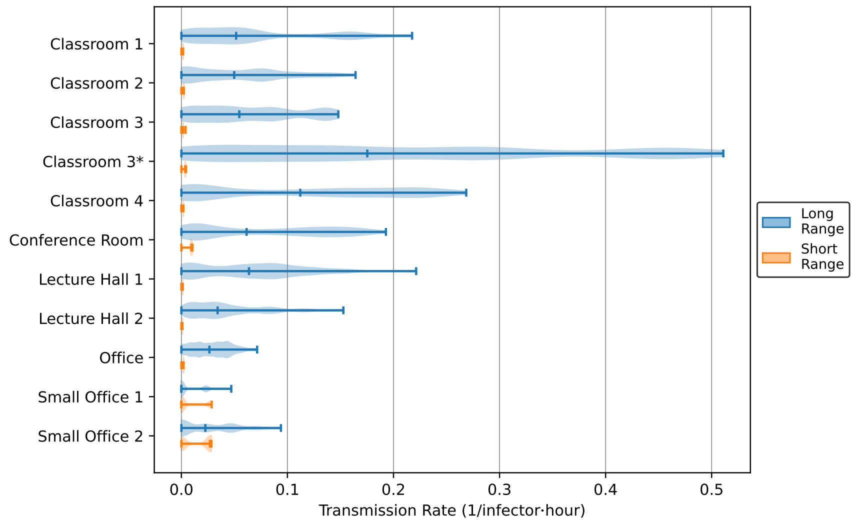

Figure 23: Distributions of short-range and long-range transmission rates in monitored spaces. Short-range rates are calculated from an empirical model based on the results of [9], while long-range rates use the pseudo-steady model from the main paper. As in Figure 5, the distributions are weighted based on the number of occupants in the space at each time point.

in line with the "Medium" scenario reported in [12] and does not violate any exposure guidelines. To estimate power consumption, we assume that the lamps operate at a constant efficiency for conversion of input power to UV radiation in the relevant 222 nm band. We estimate this efficiency to be equal to 0.0055, which follows from the experiments of [11] in which a 12 W far UV lamp was found to deliver an intensity of 100 $\mu W/cm^2$ over an area of 666 $cm^2$. The power consumption in each room can thus be calculated from the imposed intensity, the floor area of the room, and this efficiency factor.

#### 5.2 Demand-Controlled Ventilation

In the demand-controlled ventilation scenarios, time-varying ventilation is determined by standard proportional-integral control applied to the measured $CO_2$ concentration. Given time-varying bounds $Q_a^{\min}[t]$, $Q_a^{\max}[t]$, and $Q_a^{\text{mid}} := (Q_a^{\min}[t] + Q_a^{\max}[t])/2$, as well as a (possibly time-varying) $CO_2$ setpoint $C_{CO_2}^{\text{sp}}[t]$, a controller gain $k_c$, and an integral time $\tau_i = 5$ minutes, the time-varying ventilation $Q_a[t]$ is calculated in discrete time (with timestep $\delta$) as follows:

$$
\begin{aligned}
\epsilon[t] &:= C_{CO_2}[t] - C_{CO_2}^{\text{sp}}[t] \\
Q_a[t] &= \text{clip}\Big(Q_a^{\text{mid}}[t] + k_c(\epsilon[t] + e[t]/\tau_i), Q_a^{\min}[t], Q_a^{\max}\Big) \\
e[t+1] &:= \begin{cases} e[t] + \epsilon[t]\delta & \text{if } Q_a^{\min}[t] < Q_a[t] < Q_a^{\max}[t] \\ e[t] & \text{else} \end{cases}
\end{aligned}
$$

Note that the new state variable $e[t]$ represents the integral of tracking error, which is used to ensure that the $CO_2$ concentration reaches its setpoint (or ventilation reaches one of its bounds).

A key input to this algorithm is the *actual* $CO_2$ concentration $C_{CO_2}$. Because this value is directly affected by ventilation, we thus have to simulate what the $CO_2$ concentration would be under the new ventilation strategy. To accomplish this, we make use of the following model for $CO_2$ concentration:

$$
\frac{dC_{CO_2}}{dt} = g_{CO_2} + \frac{Q_a}{V}(C_{CO_2,\text{OA}} - C_{CO_2})
$$

in which $g_{CO_2} := Q_b C_{CO_2,b} N_t / V$ gives the instantaneous generation rate of $CO_2$ due to occupants. In discrete time, this equation becomes

$$
C_{CO_2}[t+1] = C_{CO2}[t] \exp(-Q_a[t]\delta/V) + g_{CO_2}[t]\delta \, \text{exprel}(-Q_a[t]\delta/V)
$$

with $\text{exprel}(x) := (\exp(x) - 1)/x$ and constant sample rate $\delta = 1$ minute. Using this equation, we can thus solve for the generation rate $g_{CO_2}[t]$ at each time using the actual measurements from the monitoring period and then simulate a new trajectory with that same generation rate but different ventilation. (Note that to reduce the effect of measurement noise, we filter the raw $g_{CO_2}[t]$ sequence via a 10-minute moving average and then clip negative values to zero prior to simulating the new trajectory; these steps ensure predictions are more physically realistic, although it will overestimate the buildup of $CO_2$ during nighttime unoccupied hours.)

This strategy can be implemented by most modern BMSs provided that a measurement of $CO_2$ concentration is available. There may be various modifications or adjustments, for example integration with economizer logic to increase outdoor-air flow under appropriate outdoor conditions, or measuring $CO_2$ in multiple places and controlling the worst-case value to its setpoint. However, the general premise is the same: outdoor airflow is adjusted so as to control the $CO_2$ concentration to its setpoint. Thus, the system will automatically increase ventilation when occupants enter the room and decrease it when occupants leave.

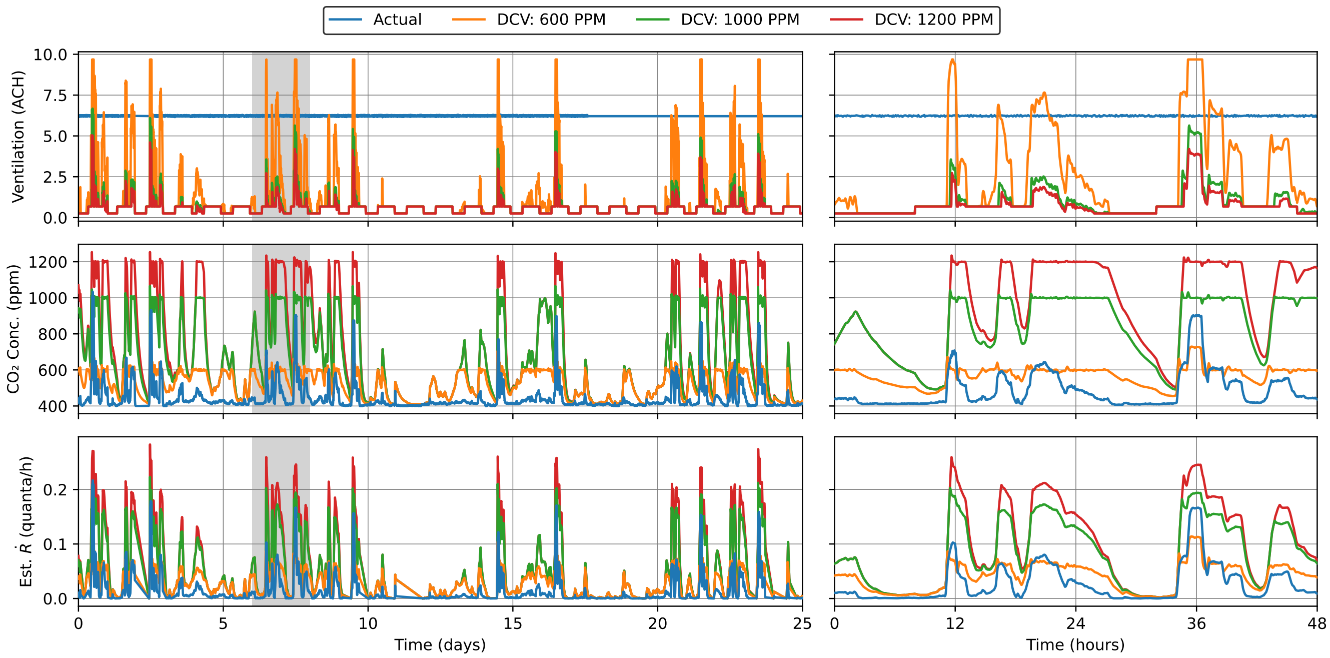

Figure 24: Simulated demand-controlled ventilation in comparison with actual operation for Classroom 1. The second column shows a zoomed-in view of the gray shaded region in the first column.

To illustrate the resulting ventilation profiles, Figure 24 shows the resulting ventilation rate, $CO_2$ concentration, and (dynamic) reproductive number for Classroom 1 as actually operated and under three different $CO_2$ setpoints. From the $CO_2$ plot, we can see that DCV does consistently achieve its control objective, keeping $CO_2$ concentration at or below its setpoint with only small overshoot (which is to be expected with this type of control following abrupt occupancy changes). However, by examining the reproductive number trajectory, we see that performance is not quite ideal: when there is a large increase in occupancy, the controller waits for the $CO_2$ measurement to exceed its setpoint before taking real action; thus, there is a period of unnecessarily high transmission risk. While this deficiency could be addressed by adjusting controller tuning or possibly adding derivative action, we instead propose using a different variable as the control objective as described in the next section.

#### 5.3 Transmission-Controlled Ventilation

Given that our primary goal is to control the transmission risk in each room, we propose using the reproductive number directly as a controlled variable, rather than using $CO_2$ as in DCV. Specifically, we propose using the *pseudo-steady* calculated transmission risk, which brings two key benefits: (1) the pseudo-steady value provides a slight degree of predictive action, as it reflects where the room is headed rather than where it currently is; and (2) the formulas are considerably simpler to evaluate.

To implement this control strategy, we of course first need to evaluate the current transmission rate $\dot{\mathcal{R}}$. The pseudo-steady model gives $\dot{\mathcal{R}} := Q_b^2 C_q N_s / \lambda_{\mathrm{EOA}} V$, in which we have removed some extra factors for brevity. The value of $\lambda_{\mathrm{EOA}} V$ can be calculated using flow measurements and filtration parameters for the BMS-provided clean air and the humidity measurements and physics-based models for the deposition and deactivation components of EOA. To estimate $N_s$, we note from the discussion of DCV that the $CO_2$ generation rate $g_{CO_2} := Q_b C_{CO_2,b} N_t / V$ can be calculated from successive measurements of $C_{CO_2}$ and $Q_a = \lambda_a V$. We could then calculate $N_i := \max(N_t - 1, 0)$, although we propose using $N_i \approx N_t$ to add a slight degree of robustness for small rooms. We thus arrive at the formula

$$\dot{\mathcal{R}} = \frac{Q_b C_q}{\lambda_{\mathrm{EOA}}} \frac{g_{\mathrm{CO_2}}}{C_{\mathrm{CO_2},b}}$$

which can be evaluated by the BMS.

To define the action of the controller, we thus take a transmission-rate setpoint $\dot{\mathcal{R}}^{\mathrm{sp}}$ and invert the previous formula to find the corresponding EOA setpoint

$$\lambda_{\mathrm{EOA}}^{\mathrm{sp}} := \frac{Q_b C_q g_{\mathrm{CO_2}}}{\dot{\mathcal{R}}^{\mathrm{sp}} C_{\mathrm{CO_2},b}}$$

From this value, the BMS can adjust its various setpoints to deliver the required amount of EOA. In cases where the BMS can control multiple sources of EOA (e.g., ventilation, filtration via recirculation, and possibly in-zone disinfection devices), some form of prioritization would be needed, for example selecting in order of increasing energy consumption. However, for the spaces being monitored, ventilation is the only directly controllable source of EOA, and thus the logic is straightforward. Specifically, we have

$$Q_a := \mathrm{clip}\Big(V(\lambda_{\mathrm{EOA}}^{\mathrm{sp}} - \lambda_s - \lambda_v - \lambda_d), Q_a^{\min}, Q_a^{\max}\Big)$$

using the sedimentation and deactivation models to calculate $\lambda_s$ and $\lambda_v$. As before, this value is clipped to (possibly time-varying) bounds to respect minimum limits and maximum capacity.

To illustrate the resulting ventilation profiles, Figure 25 shows the resulting ventilation rate, $CO_2$ concentration, and (dynamic) reproductive number for Classroom 1 as actually operated and under two different $\dot{\mathcal{R}}$ setpoints. With this modified strategy, we see that the transmission rate now stays within its established bound whenever possible (with violations occurring only when ventilation hits its upper bound). Using the pseudo-steady model allows the control system to adjust ventilation *before* the transient dynamics in $CO_2$ concentration have resolved, thus avoiding periods of unnecessarily high transmission risk.

### 6 Data Availability

All data used in this study, plus the sensor data that was collected but not used, is available at `https://github.com/acoh64/MIT-JCI-IAQ-HVAC`. The full dataset consists of timeseries data for each monitored room, one common timeseries weather file, and a table of fixed parameters for the rooms. Timeseries data files are in Parquet format, while the table of parameters is a single CSV file.

Each set of timeseries data comes in both a “raw” and “clean” version (filenames `*_raw.parquet` and `*_clean.parquet` respectively). The raw data contains values exactly as collected, including possible irregular sample rate and gaps when data collection was unavailable. The clean data is the result after interpolation and imputation to remove clear outliers and fill in missing values.

Timeseries data files for each room (filenames `<room>_*.parquet`, where `<room>` is the room name with spaces removed) contain the following columns:

- $CO_2$ Concentration (ppm)
- Occupancy (#)
- Supply Air Flow (cfm)

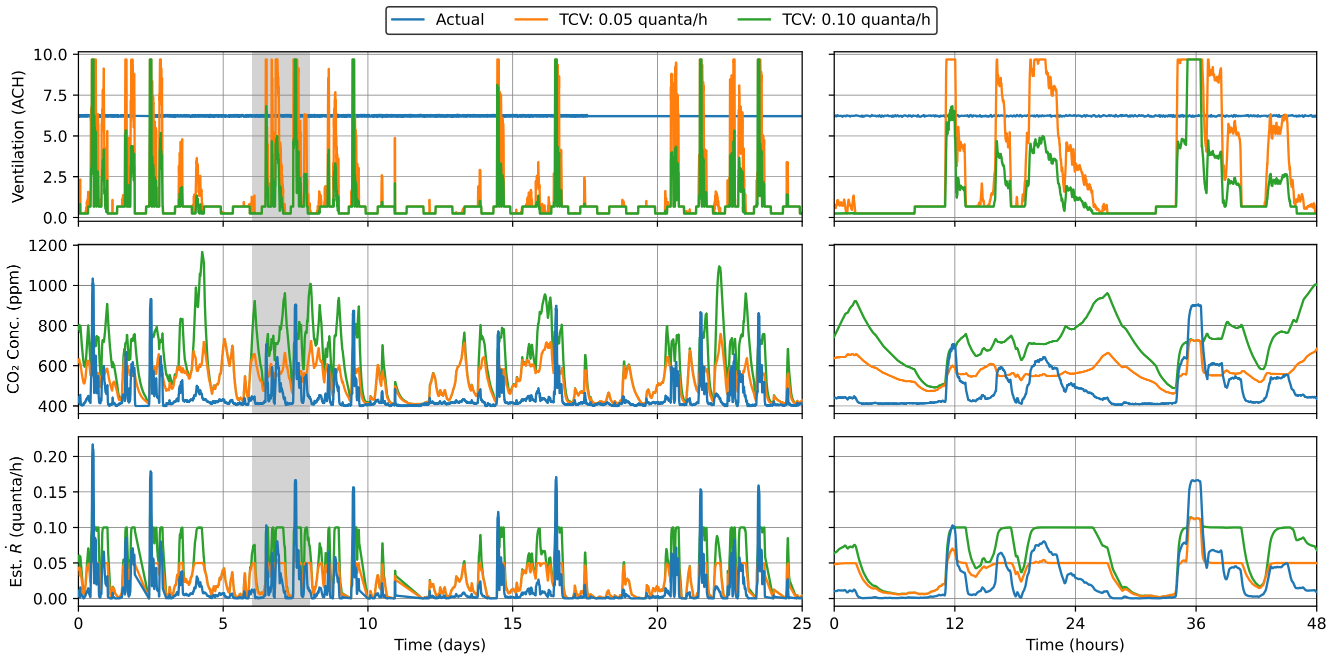

Figure 25: Simulated transmission-controlled ventilation in comparison with actual operation for Classroom 1. The second column shows a zoomed-in view of the gray shaded region in the first column.

- Supply Air Temperature (°F)
- Room Relative Humidity (%)
- Room Temperature (°F)
- TVOC Concentration (ppb)
- PM10 Concentration (μg/$m^3$)
- PM2.5 Concentration (μg/$m^3$)

Note that "TVOC" stands for "total volatile organic compounds", while "PM" is "particulate matter".

The weather data files (filenames `Weather_*.parquet`) contain the following columns:

- Outdoor Air Relative Humidity (%)
- Outdoor Air Temperature (°F)

All timeseries files contain a datetime index giving local time for each row.

The room parameter file `room_parameters.csv` contains the following columns:

- Name
- Area ($ft^2$)
- Ceiling Height (ft)
- Design Occupancy (#)
- Assumed Peak Occupancy (#)

- Mean Ventilation (ACH)
- ASHRAE Minimum Ventilation (ACH)
- Assumed Maximum Ventilation (ACH)
- In-Zone Filtration (ACH)

The “Name” column in this table correspond to the names in the timeseries data files. Values are as shown in Table 1.

#### 6.1 Timeseries Room Data Details

Timeseries room data was collected by a mix of in-room Kaiterra sensors, measurements taken by the BMS, and manually counted or imputed occupancy counts. Within each room dataset, the following columns were collected by the deployed Kaiterra sensors:

- $CO_2$ Concentration (ppm)
- Room Relative Humidity (%)
- Room Temperature (°F)
- TVOC Concentration (ppb)
- PM10 Concentration (µg/m$^3$)
- PM2.5 Concentration (µg/m$^3$)

In rooms where multiple sensors are placed, these values represent an average of the measurements. Gaps are filled by linear interpolation, with the exception of “$CO_2$ Concentration” in which large gaps were repaired with exponential decays to avoid implying significant overnight occupancy. The $CO_2$ concentration also undergoes a “recalibration” procedure so that minimum values are set to 400 ppm. (Note that the sensors themselves contain similar logic, so this procedure generally does not change values significantly. However, it was applied to remove some periods where measured values were near 300 ppm, which is unrealistically low.) Outlier removal was not otherwise performed for these data streams.

Columns collected by the BMS are as follows:

- Supply Air Flow (cfm)
- Supply Air Temperature (°F)

Note that all monitored rooms were supplied by 100% outdoor air, so the “Supply Air Flow” column gives the provided outdoor-air ventilation. Unfortunately, BMS data was not available for the final week of the study, so those periods have been imputed with constant values, as will be evident in the clean data files. Outliers from the flow measurements were removed during data cleaning. For rooms without flow measurements, values were set to the estimated ventilation rates as described in the paper. (This procedure was also applied for Office, as the measured flow includes only part of the total supply flow to the room.)

Finally, the “Occupancy” column in the clean data files are as calculated by the occupancy-estimation procedure described in the paper. We provide these values for convenience and note that alternative occupancy-estimation procedures could be applied using the other data streams. In the raw data files for Classroom 1 and Office, there are also periods where occupancy was manually counted. For other spaces, the raw occupancy data is completely empty.

#### 6.2 Timeseries Weather Data Details

To collect weather data, two QuantAQ sensors were deployed on roofs of monitored buildings. Unfortunately, these sensors were placed in areas receiving significant direct sunlight, which led to unrealistically high temperature measurements (upwards of 90 °F many afternoons). Thus, we have instead opted to use data collected from a nearby weather station. Values in the raw file are the hourly values reported from the weather station, while the values in the clean file are linearly interpolated to match the sample rate of the other timeseries data.

#### 6.3 Room Parameter Details

During sensor installation, "Ceiling Height" for each room was measured using a laser distance meter. For spaces with variable height, the value is a rough average. "Design Occupancy" was estimated by counting seats in the room. "Area" was measured from architectural drawings provided by the facilities team. "Assumed Peak Occupancy" was taken as a fraction of "Design Occupancy" based on observed room utilization. "In-Zone Filtration" was calculated from nameplate capacity of installed in-zone filtration devices (present only in Classroom 3).

"Mean Ventilation" is taken as the mean measured value in room with BMS flow measurements and set to the estimated value otherwise. "ASHRAE Minimum Ventilation" was calculated from ASHRAE Standard 62.1-2019 based on space usage. "Assumed Maximum Ventilation"" was generally estimated based on expected design heat loads, with some manual adjustments to match measured data.

### 7 Transmission Parameter Values

The parameters used in our transmission modeling and their typical ranges are given in Table 2.

| Symbol | Meaning | Typical Values |
|---|---|---|
| | **Engineering Parameters** | |
| $\tau$ | Time since an infected person entered the room | 0-1000 h |
| $V$ | Room volume | $10$–$10^4$ m$^3$ |
| $A$ | Floor surface area | 5–5,000 m$^2$ |
| $Q_a$ | Ventilation outflow rate | $1$–$10^5$ m$^3$/h |
| $\lambda_a$ | Ventilation (outdoor air exchange) rate, $Q_a/V$ | 0.1–20 h$^{-1}$ |
| $Q_r$ | Recirculation flow rate | $1$–$10^5$ m$^3$/h |
| $\lambda_r$ | Recirculation air exchange rate, $Q_r/V$ | 0.1–20 h$^{-1}$ |
| $\lambda_s$ | Drop sedimentation rate | 0.01–10 h$^{-1}$ |
| $\lambda_v$ | Pathogen (virion) deactivation rate | 0–70 h$^{-1}$ |
| $p_f$ | Probability of droplet filtration via recirculation | 0–1.0 |
| $\lambda_f$ | Filtration removal rate, $p_f\lambda_r$ | 0–30 h$^{-1}$ |
| $Q_d$ | Volumetric flow through disinfeciton device | $1$–$10^4$ m$^3$/h |
| $p_d$ | Probability of removal/deactivation in disinfection device | 0.5–1 |
| $\lambda_d$ | Total removal/deactivation rate for disinfection devices | 1–100 h$^{-1}$ |
| $\lambda_{\text{EOA}}$ | Equivalent outdoor air supply rate, $\lambda_a + \lambda_f + \lambda_s + \lambda_v + \lambda_d$ | 0.1–250 h$^{-1}$ |
| $p_m$ | Mask penetration probability | 0.01–0.1 |
| | **Physical Parameters** | |
| $r$ | Respiratory drop radius | 0.1–100 $\mu$m |
| $V_d$ | Drop volume, $\approx \frac{4}{3}\pi r^3$ | $10^{-5}$–$10^6 \mu$m$^3$ |
| $n_d$ | Drop number density per radius | 0.01–1.0 (cm$^3 \mu$m)$^{-1}$ |
| $v_s$ | Drop settling speed | $10^{-5}$–$10^2$ mm/s |
| $\lambda_s$ | Drop settling rate, $v_s(r)/H$ | $10^{-5}$–$10^2$ h$^{-1}$ |
| $x$ | Distance from infected person | 0.1-10 m |
| $p_{\text{short}}$ | Probability of being in the respiratory jet of an infected person | 0–1 |
| $Q_b$ | Mean breathing flow rate | 0.5–3.0 m$^3$/h |
| $C_{\text{CO}_2,\text{OA}}$ | Background $CO_2$ concentration | 350–450 ppm |
| $C_{\text{CO}_2,b}$ | Exhaled $CO_2$ concentration | 35,000–40,000 ppm |
| | **Epidemiological Parameters** | |
| $N_t, N_s, N_i$ | Number of total, susceptible, and infected persons | 1-1000 |
| $\beta_a$ | Airborne transmission rate per infected-susceptible pair | $10^{-6}$–10 quanta/h$^{-1}$ |
| $\bar{r}$ | Effective infectious drop radius | 0.3–5$\mu$m |
| $P$ | Pathogen production rate / air volume / drop radius | $10^{-6}$–$10^9$ (h$\mu$m)$^{-1}$ |
| $C$ | Infectious pathogen concentration / air volume / radius | $10^{-8}$–$10^4$ (m$^3\mu$m)$^{-1}$ |
| $C_s$ | Steady-state airborne pathogen concentrations, $P/(\lambda_{\text{EOA}}V)$ | $10^{-8}$–$10^4$ virions/(m$^3 \cdot \mu$m) |
| $c_v$ | Pathogen (virion) concentration per drop volume | $10^4$–$10^{11}$ RNA copies/mL |
| $c_i$ | Pathogen infectivity, 1/(infectious dose) | 0.001–1.0 |
| $C_q$ | Infectiousness of breath, exhaled quanta concentration | 1–1000 quanta/m$^3$ |
| $\mathcal{R}_{\text{in}}$ | Indoor reproductive number | 0.001–100 |
| $\epsilon$ | Risk tolerance, bound on $\mathcal{R}_{in}$ | 0.005–0.5 |

Table 2: Parameters used in transmission modeling theory with typical ranges and units.
